# Individualized fMRI connectivity defines signatures of antidepressant and placebo responses in major depression

**DOI:** 10.1101/2022.09.12.22279659

**Authors:** Kanhao Zhao, Hua Xie, Gregory A. Fonzo, Xiaoyu Tong, Nancy Carlisle, Matthieu Chidharom, Amit Etkin, Yu Zhang

## Abstract

Though sertraline is commonly prescribed in patients with major depressive disorder (MDD), its superiority over placebo is only marginal. This is in part due to the neurobiological heterogeneity of the individuals. Characterizing individual-unique functional architecture of the brain may help better dissect the heterogeneity, thereby defining treatment-predictive signatures to guide personalized medication. In this study, we investigate whether individualized brain functional connectivity (FC) can define more predictable signatures of antidepressant and placebo treatment in MDD. The data used in the present work were collected by the Establishing Moderators and Biosignatures of Antidepressant Response in Clinical Care (EMBARC) study. Patients (N=296) was randomly assigned to antidepressant sertraline or placebo double-blind treatment for 8 weeks. The whole-brain FC networks were constructed from pre-treatment resting-state functional magnetic resonance imaging (rs-fMRI) at 4 clinical sites. Then, FC was individualized by removing the common components from the raw baseline FC to train regression-based connectivity predictive models. With individualized FC features, the established prediction models successfully identified signatures that explained 22% variance for the sertraline group and 31% variance for the placebo group in predicting HAMD17 change. Compared with the raw FC-based models, the individualized FC-defined signatures significantly improved the prediction performance, as confirmed by the 10×10-fold cross-validation. For sertraline treatment, predictive FC metrics were predominantly located in the left middle temporal cortex (MTC) and right insula. For placebo, predictive FC metrics were primarily located in the bilateral cingulate cortex and left superior temporal cortex (STC). Our findings demonstrated that individualization of FC metrics through removal of common FC components enhanced the prediction performance compared to the raw FC. Associated with previous MDD clinical studies, our identified predictive biomarkers provided new insights into the neuropathology of antidepressant and placebo treatment.

## Introduction

Major depressive disorder (MDD) is one of the most prevalent and recurrent mental illnesses, impacting millions of people worldwide.^1^ Antidepressants such as selective serotonin reuptake inhibitors (SSRIs), have been the treatment cornerstone for MDD since the 1960s,^2^ yet they show efficacy in only about 57% of patients,^3^ with slightly higher efficacy than placebo (Cohen’s d of ∼0.3).^3, 4^ Uncovering the reason why MDD patients respond heterogeneously^5^ to antidepressants is critical to grounding our understanding of the underlying neurobiological mechanisms of these treatments, thus providing directions for the development of novel antidepressant medications.

Over the past decades, non-invasive neuroimaging techniques, such as functional magnetic resonance (fMRI) and electroencephalography, have been increasingly applied to probe brain dysfunction of various mental disorders.^6, 7^ Moreover, resting state functional connectivity (FC), characterizing the synchronized intrinsic function of brain regions across time,^8^ offers new insights to inform understanding of neural communication and cognitive processes.^9^ Previous studies have successfully identified depression-relevant brain dysfunctions by FC analysis.^10, 11^ For example, default mode network (DMN) dysfunction, particularly within the anterior cingulate cortex and medial prefrontal cortex, has been implicated in MDD.^12, 13^ In recent years, an increasing number of studies have begun to discover promising FC biomarkers predictive of treatment response in MDD.^14^ FCs involved in anterior cingulate cortex and prefrontal cortex were shown to predict treatment remission of antidepressant medication.^15-17^ Additionally, pre-treatment FC between the dorsolateral prefrontal cortex and insula showed a significant correlation with symptom improvement in MDD after treatment by repetitive transcranial magnetic stimulation.^18^

The human cerebral cortex is typically organized into several distributed distinct function networks.^19^ Emerging studies have demonstrated the individual uniqueness in subtle and reliable change of functional network architecture and in potential clinical application.^20-22^ For instance, the individualized FC were obtained by extracting the FC from iteratively refined individual-specific parcellation of each subject^20, 23, 24^. Based on individualized brain functional organization, the sensitivity of brain stimulation therapy has been improved.^20^ Moreover, recent studies have utilized individual-specific brain functional connectivity to investigate the neural correlates of clinical measurements.^24^ The quantified individual-unique FC features could more precisely predict the symptom severity of various psychotic illnesses.^23, 24^ Similarly, common components of fMRI blood-oxygenation-level-dependent (BOLD) signals shared across subjects were found to obfuscate the prediction model, leading to lower prediction performance for personality and emotion measures.^25^ Nevertheless, to the best of our knowledge, no existing studies have investigated the efficacy of using individualized FC to predict antidepressant outcomes. We hypothesized that by removing the common components, individualized FC would better capture the depression treatment-related dimensions to increase treatment outcome prediction performance. The predictive neural circuits driven by individualized FC may further facilitate the understanding of neurophysiological mechanisms of antidepressants. Furthermore, typical FC individualization methods^20, 23, 24^ iteratively refined individual-specific parcellation for each subject based on voxel level time series. Compared with typical individualization method, our method required less computation resource without calculation of individualized parcellation for each subject.

By leveraging data from the Establishing Moderators and Biosignatures of Antidepressant Response in Clinical Care (EMBARC) study, a large multi-site trial examining predictors of response to sertraline (belongs to the group of SSRIs) vs. placebo treatment in major depression,^26^ we developed a machine learning framework for modeling individualized FC and examined its efficacy in quantifying treatment-predictive biomarkers in MDDs. We first utilized an unsupervised group component analysis approach to extract individualized FC by removing common connectivity components shared across subjects. We then built a sparse regression model based on the individualized FC features (residual features remaining after removal of common components) for predicting individual-level treatment outcomes. The efficacy of the prediction was cross-validated on sertraline and placebo treatment arms, respectively. We investigated the superiority of using individualized FC versus raw FC for outcome prediction and to interpret how individualization improved FC predictive validity. Linear mixed effect analysis was further conducted to confirm the capacity of the identified individualized FC metrics for treatment outcome prediction under a longitudinal intent-to-treat framework.

## Methods

### Clinical trial data

In this study, we leveraged data from the EMBARC study,^26^ the largest neuroimaging-coupled placebo-controlled randomized clinical trial of depression to date. Eligible patients (ages 18 - 65) were recruited at four study sites (Massachusetts General Hospital, University of Texas Southwestern Medical Center, University of Michigan, and Columbia University). Employing a double-blind randomized controlled trial design, patients with a diagnosis of MDD were assigned to 8 weeks of placebo or sertraline. More details about the inclusion criteria can be found in supplement section M1 and prior publications.^26^ Of 309 patients diagnosed with MDD, 296 subjects who passed the inclusion criteria received either placebo or sertraline treatment. The severity of depression symptoms was assessed with the 17-item *Hamilton Rating Scale for Depression*^27^ (HAMD_17_) at each study visit (baseline, weeks 1, 2, 3, 4, 5, 6, and 8). The missing HAMD_17_ scores at week 8 were imputed using Bayesian regression.^16^ Treatment outcome was quantified as the difference in HAMD_17_ score from baseline to week 8.

### Functional MRI acquisition and preprocessing

The acquired rs-fMRI data were preprocessed using the reproducible fMRIPrep pipeline (see supplement M1 for more details).^28^ The ones with low signal quality, such as a large framewise displacement value, were excluded from the subsequent analysis. Finally, we obtained usable rs-fMRI data from 131 patients with sertraline treatment and 144 patients with placebo treatment. Patients in these two arms showed no significant differences in the sociodemographic and clinical variables (Figure S4).

### Functional connectivity feature calculation

The preprocessed voxel-level BOLD signals were averaged into 100 region-of-interest (ROI) level time series according to the Schaefer et al. parcellation.^29^ For each subject, raw FC was obtained by computing the Pearson’s correlation coefficient of time series between any pair of ROIs. Fisher’s r-to-z transformation was applied to ensure the normality of the connectivity features, followed by z-score normalization.

### Extraction of individualized functional connectivity

To calculate individualized FC, we utilized common orthogonal basis extraction (COBE), a powerful algorithm for common and individual feature extraction from multi-set data.^30^ Assuming 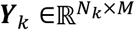 (*k* ***=*** *1,2,…K*; *N*_*k*_, the number of subject in *k****-***th group; *M*, number of FC) are the FC matrices of *K* groups of subjects, COBE aims to decompose these FC matrices into common features shared across subjects and individual-specific features:

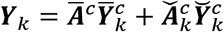

where ***Ā***^*c*^ represent the loading corresponding to the *c*-th common component, 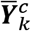 represent the corresponding coefficient matrix of *k*-th grouped data, and 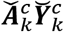 represent projection of the *k*-th subjects in individual subspace. More theoretical background can be found in prior work^30^. The illustration of COBE-based FC individualization was provided in Figure S1. Individualized FC was obtained via removing the reconstructed common connectivity from raw FC to characterize individual-specific (residualized) brain functional architecture. Consistent with previous literature^31, 32^, the hyperparameter *K* was set to 10 in our analysis. The number of common components *C* was set to 5 since the first five components together explained near 70% variance (Figure S5A). More details about the discussion of hyperparameter (*K* and *C*) optimization in COBE was provided in Supplemental M4.

### Connectome predictive modeling of treatment response

With the individualized FCs, we combined connectome predictive modeling (CPM) and Lasso regression to build machine learning models for predicting treatment response. In the CPM step, FC metrics showing significant correlation to the treatment outcome were selected as informative features for the prediction model training. Furthermore, Lasso regression, a typical machine learning technique with a sparsity constraint on the feature dimension, was applied to refine the feature set and build a robust prediction model. The prediction performance was evaluated by 10×10-fold cross-validations (CVs). The outcome prediction performance was evaluated by computing Pearson’s correlation coefficient and R-squared value between the predicted HAMD_17_ score changes and the observed ones. False discovery rate (FDR) correction was used to correct the statistical significance (*p* value) across all treatment predicted models. More details about the implementation of CPM and LASSO-based prediction were provided in Supplemental M5, Figure S12. An overview of the analytical framework was illustrated in Figure S3.

### Effect analysis of functional connectivity individualization

To examine the effects of FC individualization on improving treatment outcome prediction, Wilcoxon signed-rank test was used to compare *R*^*2*^ values and Pearson correlation coefficients of ten repeated runs of 10-fold CVs of individualized FCs and raw FCs. To explore why individualized FCs enhanced the prediction performance, we first obtained the significantly changed weights in the Lasso model trained using individualized FCs versus raw FCs, across 100 folds, via Wilcoxon signed-rank test. Second, the change of Pearson’s correlation coefficient between the HAMD_17_ change and predictive individualized FCs in the CPM step, was evaluated by the Wilcoxon signed-rank test. Finally, the critical changed connectivity was defined as the pairwise FCs with significantly changed prediction weight as well as with significantly changed correlation coefficient to HAMD_17_ change, pattern post-FC-individualization. (see supplement M6 for more details)

### Linear mixed effect analysis

To examine how the informative FC patterns identified by the prediction models were correlated with the trajectory of HAMD_17_ change across study visits, linear mixed effect models (LME) in an intent-to-treat analysis were applied. Specifically, the dependent variable was HAMD_17_ at each study visit, and the independent variable was individualized FC strength label (binarize all subjects into low or high categories, thresholding based on the median of the FC), time, and FC strength by time interaction. We focused on a significant FC-by-time interaction, indicating how the FC value was associated with the HAMD_17_ change across eight weeks. All *p* values of interaction effect were FDR corrected across all predictive pairwise FCs revealed from our prediction models.

### Leave-study-site-out analysis

To verify the model generalizability to unseen data acquired using various MRI scanners in different study sites, we conducted leave-study-site-out cross-validation. We assumed that for leave-study-site out analysis, the site effect was vital for a good performance and normalization might not be enough to alleviate the site effect. Thus, we used ComBat, a powerful site-effect correction method,^33^ to harmonize the FCs across different sites based on patients’ gender, age, education, and race information. Then, the data from three study sites was iteratively used to train the model, and that from the rest site was used to test the model.

## Results

### Individualized functional connectivity improves treatment outcome prediction

For the sertraline arm, the true HAMD_17_ score change correlated significantly with the predicted HAMD_17_ score change derived from the model training with both the individualized FCs (Figure 1A, Pearson’s *r* = 0.47, *p*_*fdr*_ = 2.8 × 10^−8^, *R*^*2*^ = 0.22) and the raw FCs (Figure 1B, raw FCs, Pearson’s *r* = 0.41, *p*_*fdr*_ = 1.7 × 10^−6^; *R*^*2*^ = 0.17), but not the common FCs (Figure S10A, common FCs, Pearson’s *r* = -0.27, *p*_*permute*_ = 0.99; *R*^*2*^ = -0.04). Critically, the individualized FCs significantly outperformed raw FCs and then outperformed common FCs, as evaluated from the Wilcoxon signed-rank test of their performance (Figure S16A, for *R2, w*_*individualized vs raw*_ = 2.57, *p*_*individualized vs raw*_ = 0.014; *w*_*raw vs common*_ = 3.78, *p*_*raw vs common*_ = 0.001; Figure S16C, for *r, w*_*individualized vs raw*_ = 2.72, *p*_*individualized vs raw*_ = 0.014; *w*_*raw vs common*_ = 3.78, *p*_*raw vs common*_ = 0.001). The leave-study-site-out analysis showed the generalizability of the individualized FC-based prediction model to an unseen study (Figure S14A, with Combat correction, Pearson’s *r* = 0.42, *p*_*fdr*_ = 8.0 × 10^−7^; *R*^*2*^ = 0.11. Figure S14C, with z-score normalization, Pearson’s *r* = 0.33, *p*_*fdr*_ = 1.1 × 10^−4^; *R*^*2*^ = 0.10). For both the models trained with individualized FCs and raw FCs on the sertraline arm, applying them to the placebo arm failed to predict treatment outcome (Figure S9A, B, *r*_*individualized*_ = 0.00, *r*_*raw*_ = 0.04, *p*_*individualized*_ = 0.97, *p*_*raw*_ = 0.77). This cross-arm prediction analysis demonstrated the specificity of the models for sertraline efficacy prediction. The top 20 important FCs for the prediction of sertraline treatment outcomes were displayed in Figure 1C. The more strongly predictive FC metrics with larger weights were found between the left middle temporal cortex (MTC) in visual network (VIS) and the right supramarginal gyrus, as well as between the right inferior frontal cortex and the right insula. Again, as shown in the node strength plot (Figure 1D), the most important hubs (a node with a larger connected degree above average degree of whole FC pattern) were dominant across the right insula, left MTC, and right supramarginal cortex.

**Figure 1.**
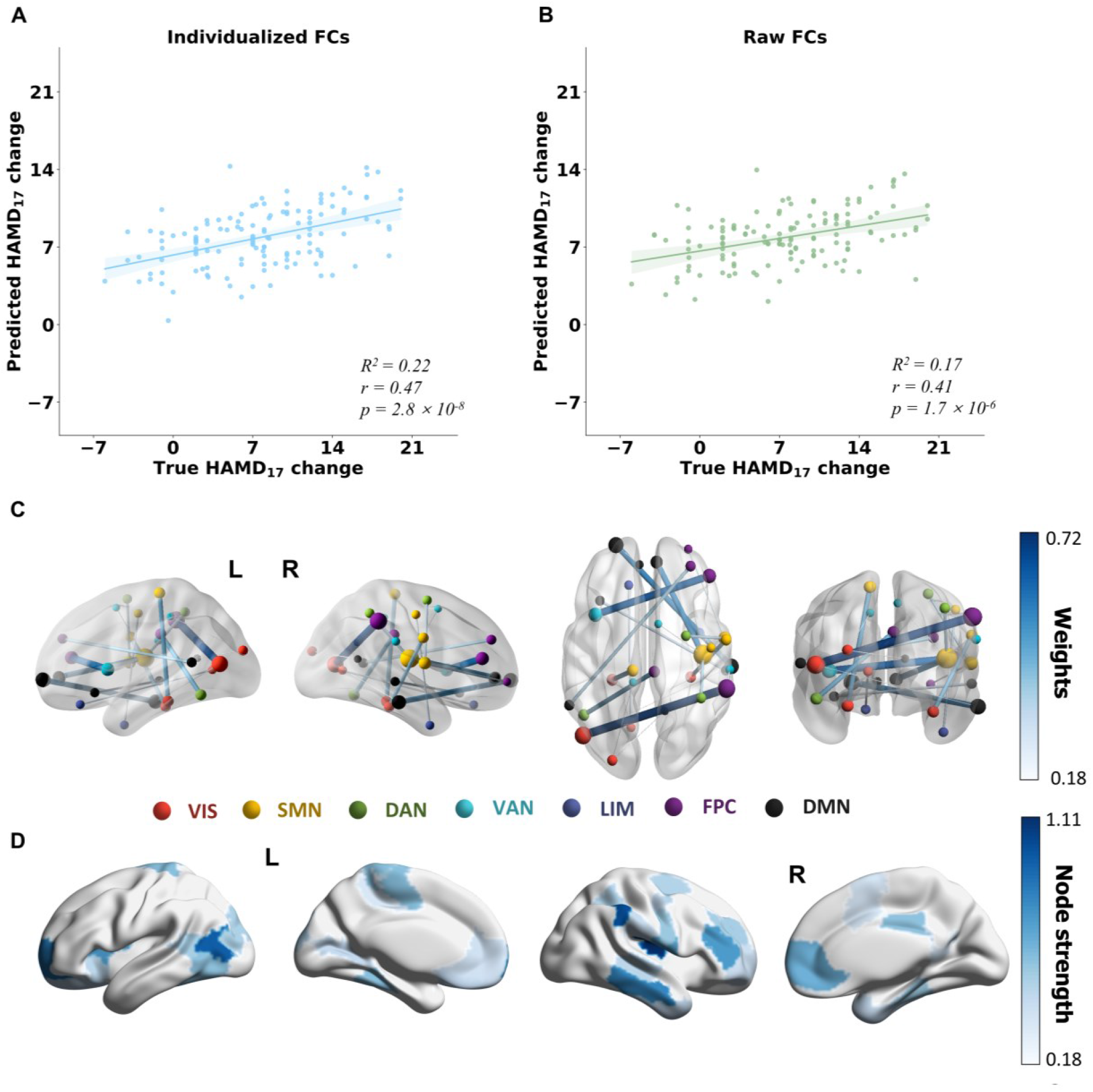
Prediction of the outcome specific to sertraline arm in 10 × 10 cross-validations. All p values were FDR corrected. **A** Prediction of HAMD_17_ change using the individualized FCs-based model (R^2^ = 0.22, Pearson’s r = 0.47, p = 2.8 × 10^−8^ based on the one-sided test against the alternative hypothesis that r > 0). **B** Prediction of HAMD_17_ change using the raw FCs-based model (R^2^ = 0.17, Pearson’s r = 0.41, p = 1.7 × 10^−6^ based on the one-sided test against the alternative hypothesis that r > 0). **C** Connectivity pattern corresponding to the top 20 model weights derived with individualized FCs. Deeper blue and thicker links represent larger model weights (absolute value). **D** Regional importance (node strength) was calculated based on the top 20 predictive connections shown in **C**. VIS, visual network; SMN, somatomotor network; DAN, dorsal attention network; VAN, ventral attention network; LIM, limbic network; FPC, frontoparietal control network; DMN, default mode network.

For the placebo arm, the true HAMD_17_ score change correlated significantly with the predicted HAMD_17_ score change derived from the model training with both the individualized FCs (Figure 2A, Pearson’s *r* = 0.56, *p*_*fdr*_ = 3.5 × 10^−12^, *R*^*2*^ = 0.31), and the raw FCs (Figure 2B, raw FCs, Pearson’s *r* = 0.48, *p*_*fdr*_ = 3.9 × 10^−9^; *R*^*2*^ = 0.23) but not the common FCs (Figure S10B, common FCs, Pearson’s *r* = -0.18, *p*_*permute*_ = 0.782; *R*^*2*^ = -0.04). The individualized FCs significantly outperformed raw FCs and then outperformed common FCs, as evaluated from the Wilcoxon signed-rank test of their performance (Figure S16 B, D, Wilcoxon signed-rank test result, for *R*^*2*^, *w*_*individualized vs raw*_ = 3.02, *p*_*individualized vs raw*_ = 0.006; *w*_*raw vs common*_ = 3.78, *p*_*raw vs common*_ = 0.001, for *r, w*_*individualized vs raw*_ = 3.18, *p*_*individualized vs raw*_ = 0.004; *w*_*raw vs common*_ = 3.78, *p*_*raw vs common*_ = 0.001). The leave-study-site-out cross-validation demonstrated the generalizability of our models to data collected from unseen study sites (Figure S14B, with Combat correction, Pearson’s *r* = 0.36, *p*_*fdr*_ = 8.6 × 10^−6^, *R*^*2*^ = 0.10. Figure S14D, with z-score normalization, Pearson’s *r* = 0.31, *p*_*fdr*_ = 1.3 × 10^−4^; *R*^*2*^ = 0.09). The cross-arm prediction analysis demonstrated the specificity of the models for placebo efficacy prediction (Figure S9A, B, *r*_*individualized*_ = 0.08, *r*_*raw*_ = 0.07, *p*_*individualized*_ = 0.55, *p*_*raw*_ = 0.55). For the placebo arm (Figure 2C, D), the most important hubs were mainly located in the bilateral middle cingulate cortex, left superior temporal cortex (STC), left posterior cingulate cortex (PCC), and the right orbital frontal cortex. For both sertraline and placebo treatment, the clinical measurements listed in Supplement M3 except HAMD_17_ could not predict the outcome (Figure S15).

**Figure 2.**
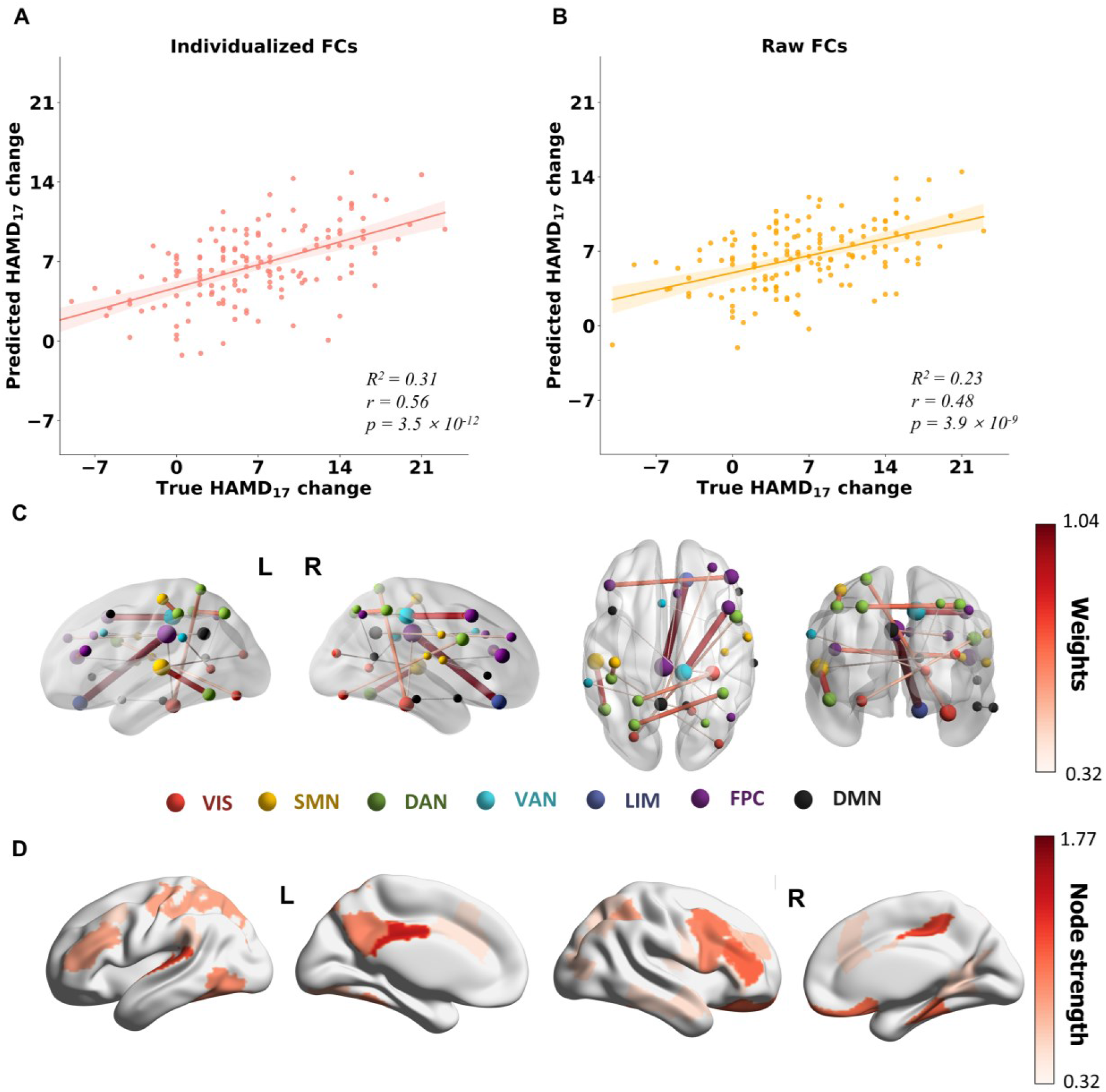
Prediction of the outcome specific to placebo arm in 10 × 10 cross-validations. All p values were FDR corrected. **A** Prediction of HAMD_17_ change using the individualized FCs-based model (R^2^ = 0.31, Pearson’s r = 0.56, p = 3.5 × 10^−12^ based on the one-sided test against the alternative hypothesis that r > 0). **B** Prediction of HAMD_17_ change using the raw FCs-based model (R^2^ = 0.23, Pearson’s r = 0.48, p = 3.9 × 10^−8^ based on the one-sided test against the alternative hypothesis that r > 0). **C** Connectivity pattern corresponding to the top 20 model weights derived with individualized FCs. Deeper red and thicker links represent larger model weights (absolute value). **D** Regional importance (node strength) was calculated based on the top 20 predictive connections shown in **C**.

### Common connectivity components extracted by COBE

To identify neural circuits that contributed most strongly to the common components, the top 40 connections in the first five-component loading matrices were visualized (Figure S6). Pearson’s correlation between each component-projected features and various clinical measurement was calculated to explore the clinical/behavioral association of each common component. The first common component was mainly driven by connections between DMN and ventral attention network (VAN), and between DMN and dorsal attention network (DAN). The second component primarily consists of edges from the VIS. For the third common component, the most predominant connections were involved in the SMN and the middle and lateral temporal cortex of DMN. The fourth component was dominated by the connections from the inferior and middle frontal gyrus in FPC to SMN. Additionally, as indicated in Table S1, the first component had a positive correlation with the neuroticism score of Neuroticism-Extraversion-Openness inventory (NEO) and the general distress severity in the mood and anxiety symptom questionnaire (MASQ). The second component was positively correlated to the extraversion score of NEO and the anxious arousal frequency in MASQ. The third component was positively correlated to the anhedonic depression in MASQ and negatively correlated to the extraversion score of NEO.

To explore the summarized common effect from all components, we reconstructed common FCs for all MDDs via the five components. Then, the associations of the features projected based on the five common components with various clinical measurements were verified by multiple linear regression. The top 40 reconstructed connections, averaged across all patients, were shown in Figure S8. The hubs were middle frontal cortex in DMN, supplementary motor area in VAN and calcarine, lingual cortex from VIS. The common effect was associated with the three personality dimensions such as neuroticism, openness, and agreeableness (Table S2).

### Significantly changed connectivity pattern post FC individualization

After individualization, the FCs showing significant changes in predictive weights and correlation strengths with outcomes were shown in Figure 3. Moreover, regions showing the most critical changed connectivity patterns partly overlapped with the regions of top 20 predictive connectivity patterns driven by the Lasso regression model (blue links in Figure 3A (1) and red links in Figure 3B (1)). We observed that the overlapping top weights in critical changed pattern in our models were significantly different from the chance models (Dice coefficient was used to evaluate the degree of overlapping. Dice_*Sertraline*_ = 0.20, *p*_*permute*_ = 9.9 × 10^−4^; Dice_*Placebo*_ = 0.48, *p*_*permute*_ = 9.9 × 10^−4^). In other words, post-FC individualization, part of the critical changed FCs contributed greatly for the model prediction power. For the sertraline-predicting models, the critical changed FC patterns were involved in the right insula, right inferior temporal cortex, left MTC and left postcentral cortex (Figure 3A). The critical changed connectivity weights of the placebo predictive model were from the left STC, bilateral inferior frontal cortex, and right middle cingulate cortex (Figure 3B).

**Figure 3.**
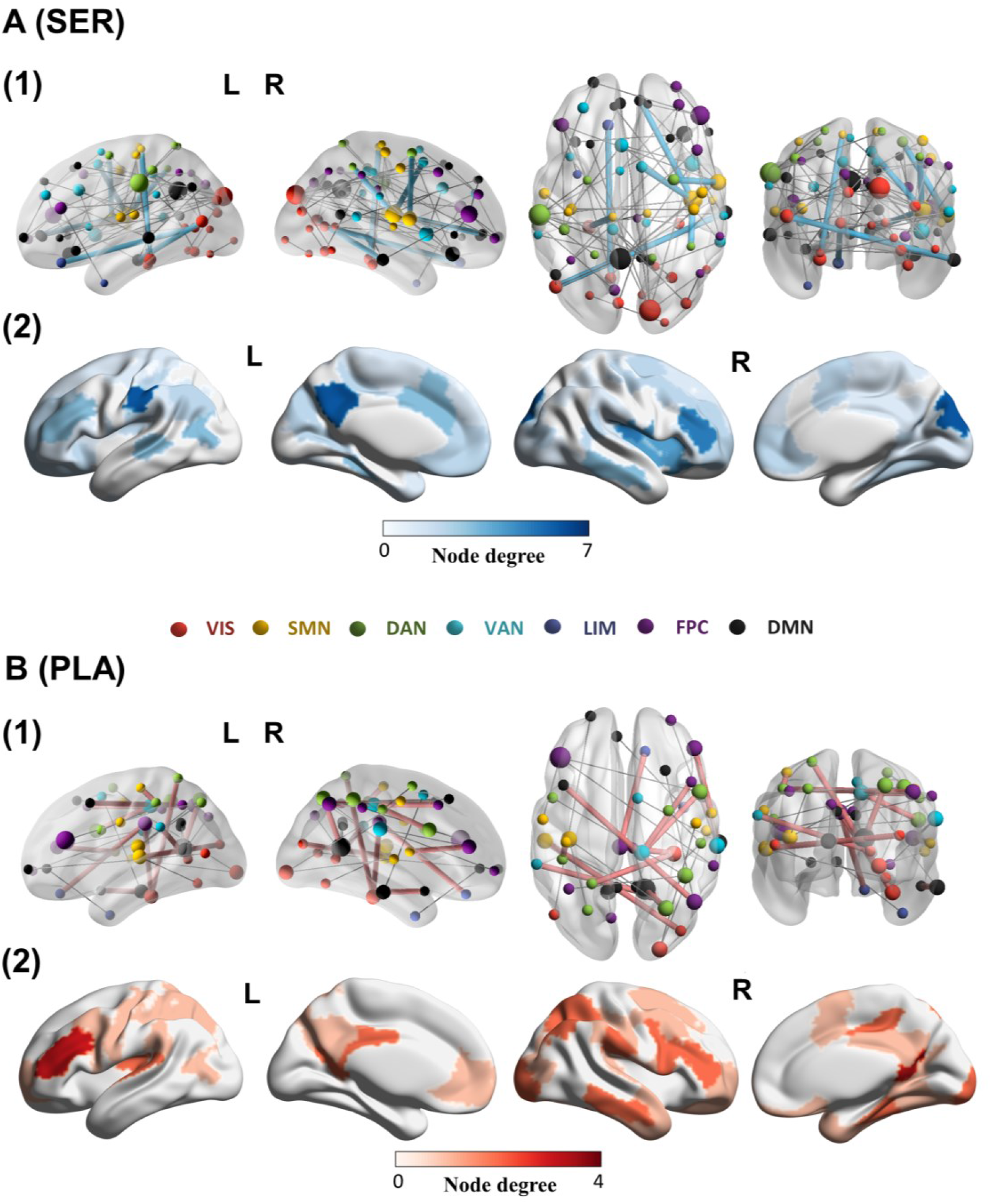
The visualization of the critical changed connectivity patterns after FCs individualization. The critical changed FCs were defined as the ones that had a significantly changed correlation to HAMD_17_ score change and significantly changed predictive pattern weights post FC individualization, evaluated from the Wilcoxon signed-rank test. Brain glass plot of the significantly critical changed pairwise FCs survived from FDR correction for model predictive to **A** sertraline arm and **B** placebo arm. **A** (1) The critical changed connections, which were overlapping the top 20 sertraline predictive FC patterns were amplified and colored blue, otherwise grey. (2) The degrees of brain regions involved in the critical changed connections were rendered on the cortical surface. **B** (1) The critical changed connections, which were overlapping the top 20 placebo predictive FC patterns were amplified and colored red, otherwise grey. (2) The degree of brain regions involved in the critical changed connections were rendered on the cortical surface.

### FC moderator of treatment outcome in eight weeks

In this section, we explored how the predictive FC patterns identified by the models interacted with the trajectory of HAMD_17_ change via applying LME analysis. For the sertraline arm, the lower FC between the right temporal pole and insula and the higher FC between the right postcentral cortex and left middle temporal cortex indicated a better sertraline treatment outcome (Figure S11). For the placebo arm, the lower FC between the left superior temporal cortex and middle occipital cortex and the lower FC between the anterior cingulate cortex and PCC suggested a better placebo treatment outcome (Figure S12). All FCs that significantly predicted treatment effects in eight weeks were summarized in Tables S3 and S4.

## Discussion

In this study, we built novel individualized FC-based machine learning models to quantify biomarkers of treatment responses to antidepressant sertraline versus placebo in major depression. Our comprehensive analyses demonstrated that the individualized FCs led to a discovery of robust signatures for the individual-level treatment response to antidepressant sertraline and placebo. Additionally, the generalizability of individualized FC signatures was confirmed by leave-study-site-out analyses. Our biomarker findings further advance the understanding of the relationship between whole-brain neural circuits and the effects of antidepressant medication and placebo. Taken together, these findings demonstrate the utility of this novel analytic approach in isolating individual neural characteristics that most strongly predict individual treatment outcomes to antidepressants, which paves the way for a potentially improved capacity to personalize psychiatric treatments.

### Shared mechanisms in antidepressant and placebo treatment effects

An interesting finding of our study showed that the right prefrontal lobe played an important role in both sertraline and placebo treatment outcome prediction even though the FCs linked from this hub were different between the two arms. In LME analysis results (Tables S4, S5), higher FC between right frontal lobe and right rolandic operculum indicated a better sertraline response and higher FC within the middle frontal cortex indicated a better placebo response. The associations of middle frontal cortex dysfunctional activation with depression symptoms have been reported in previous literature.^34, 35^ For example, lesions in the prefrontal cortex served as a risk indicator for more severe depression symptoms.^35^ The function of right middle frontal cortex was related to the maintenance of mood state, emotion perception, and behavioral control in MDD.^36^ In addition, most of the hubs of predictive FC specific for sertraline treatment outcomes, e.g., the left MTC and insula, and those specific for placebo treatment outcome, e.g., the left STC and cingulate cortex, play vital roles in regulating emotion and action and processing perceived information.^37-40^ Thus, we speculate that sertraline and placebo may both serve to regulate the participants’ emotions and action control via altering the function of both treatment-specific brain regions and common overlapping regions, which may serve as one mechanism to alleviate depression. Additional indirect evidence is that previous studies have found overlapping activated endogenous opioid neurotransmission and changes in regional glucose metabolism for both antidepressant effect placebo treatment.^41, 42^

### Neural predictors specific for antidepressant and placebo treatment

Our findings identified that FC metrics involving the left middle temporal cortex and the right insula were critical for the sertraline treatment prediction model. These FCs and corresponding glucose metabolism reported in previous studies were associated with depression-relevant brain dysfunctions^11, 36^ and antidepressant effect^43, 44^. For the MTC region, stronger functional connectivity was observed in MDDs compared with healthy individuals, and metabolic rate was found to positively correlate with the improvement of depressive symptoms.^11, 43^ Greater oxygenated-hemoglobin activation during a verbal fluency task in the left MTC at baseline correlated with a greater HAMD_17_ decrease after antidepressant treatment.^45^ This is in line with our LME result, which found that higher FC between postcentral cortex and MTC was associated with a better sertraline response. For the insula, our LME result indicated that lower FC with the right temporal pole predicted a better sertraline response. Previous study reported that decreased functional connectivity in older depressed individuals was related to apathy symptoms.^46^ The SSRI treatment effect worked in attenuating the activation of the insula during processing of emotional faces.^47^ Thus, we hypothesized that the antidepressant effect might be related to the function of emotional processing and control of MTC and insula.^37, 38, 48^

In the placebo specially-predictive FC pattern, the dominant ROI hubs were mostly located in the left superior temporal cortex (STC) and cingulate cortex. As revealed from LME analysis results, the lower connectivity between STC and left middle occipital cortex, between the anterior cingulate cortex and posterior cingulate cortex suggested a better placebo response. The dysfunction of STC and cingulate cortex in emotional processing, social cognition, and sensation regulation^39, 40, 49^ might be implicated in pathophysiology of MDD. During the face emotion evaluation task, a significant activation difference in STC of MDDs who accepted placebo and SSRI treatment was detected.^50^ The antidepressant-resistant depression patients with lesions of middle cingulate cortex exhibited a dysfunction of cognitive perception and emotional information processing such as insensitivity in recognizing fear, and anger^51^. Additionally, for cingulate cortex, the change of BOLD activity is placebo-related rather than pharamacologically-induced.^41, 52^ In our cortical surface plot (Figures 1E, 2E), the cingulate connection was more dominant in placebo treatment prediction than SSRI.

Some of important brain regions revealed from predictive pattern in our study were congruent with existing study which applied LME analysis for EMBARC.^53^ The right frontal lobe in DMN was critical for sertraline and placebo response prediction. However, for their sertraline prediction, within-network connectivity of DMN was the most important prediction biomarker and hippocampus was reported as key region for prediction of both treatment responses. In our sertraline predictive pattern, the FCs between DMN and SMN (e.g. FCs between posterior cingulate cortex and precentral cortex, between postcentral cortex and MTC) were with larger weight. The hippocampus region is not included in a typical brain parcellation, which was used in our study, in future, it is worthwhile to explore how hippocampus region contributes the model prediction with individualized FC.

### Neuropathology of FC individualization

After FC individualization, the predictive FC metrics with significant weight changes showed a significantly changed correlation to the treatment outcome (Figure 3). More importantly, those critical changed FCs were partly included in the FCs with the top 20 largest weights. The brain regions with stronger node strength were also partly overlapping in the brain regions with stronger critical changed FC node degrees. Thus, the performance improvement of the individualized FC-trained model might be related to the change in the correlation between individualized FC and HAMD_17_ change. The individualization of FCs may have captured more depression change-related neural activity, which in turn improved the prediction performance.

Besides the correlation of the individualization effect to depression severity change, the five common components removed were significantly correlated to three personality dimensions evaluated by the NEO inventory (Table S1). As indicated by our results, the first component was positively correlated to neuroticism score and the third component was negatively correlated to neuroticism score. The functional connections between DMN and VAN, and between DMN and SMN were predominant in the first and third components (Figure S6). Previous studies have linked the self-generated thought generated in DMN to neuroticism^54^ and greater neuroticism was associated with stronger FCs in SMN, DMN, and VAN,^55^ which was consistent with our results. However, the opposite correlation between these two components with neuroticism scores might derive from different top connection patterns for these three networks. Moreover, the association among the important FCs in the third common component and depression, anxiety severity and neuroticism severity revealed in our work was congruent with the overlapping symptoms between neuroticism and depression, anxiety^56^.

### Limitations

Several limitations and potential extensions of the present study are important to note. Though the sample size of our dataset was larger than most of the existing studies for antidepressant treatment prediction, replication analysis with a larger sample size is needed to validate our biomarker findings.^57^ We only trained a treatment prediction model based on fMRI data at the baseline. Future studies can focus on leveraging longitudinal neuroimaging data to further delineate the relationship between FC changes and treatment effects. Moreover, extended work can explore the causality between treatment-predictive FC metrics and treatment effects. For instance, LME analysis indicated that lower FC between right temporal pole and right insula was associated with better sertraline treatment outcome. For patients with antidepressant-resistant depression, it may be possible to utilize neuromodulatory approaches to lower the FC between these regions, which might promote an enhanced response to antidepressants. This approach would help to strengthen causal inference regarding the role of FC between right temporal pole and right insula in dictating antidepressant treatment response. Finally, although we demonstrated that our FC individualization method has significantly improved the model prediction power, the R^2^ was still marginally better than raw FC model. In future, it is essential to combine our method with other individualized parcellation based FC individualization^20^ to expect a more substantial improvement of prediction efficacy.

## Conclusion

In summary, utilizing a novel machine learning framework and individualized FC, we constructed a brain connectivity metric with superior performance in quantifying treatment predictive biomarkers in individuals with MDD. This approach provided enhanced performance relative to the model trained on raw FC metrics. The model improvement derived from individualized FCs might be associated with the changed correlation between those predictive FCs and treatment response. The right insula and left middle temporal cortex were the most predictive biomarkers of the sertraline response. The left superior temporal cortex and cingulate cortex were the most predictive biomarkers of the placebo response. After FC individualization, the predictive patterns involved in those regions were further changed. These findings provide novel insights and an enhanced method of leveraging fMRI functional connectivity towards personalized treatment selection, which advances efforts toward precision medicine in major depression disorder.

## Supporting information

supplement of all results

## Data Availability

All data produced are available online at https://nda.nih.gov/edit_collection.html?id=2199

