## supplement of all results for "Individualized fMRI connectivity defines signatures of antidepressant and placebo responses in major depression"

### Contents

#### Supplementary methods

##### Section M1: Participants and treatment

In the EMBARC database, a total of 309 participants (ages 18 - 65) were recruited at four sites: Massachusetts General Hospital, University of Texas Southwestern Medical Center, University of Michigan, and Columbia University. Across all sites, participants signed informed consent and received financial compensation. Patients with MDD met the criteria of *Diagnostic and Statistical Manual of Mental Disorders, fourth edition* (DSM-IV),[1, 2] and scored ≥ 14 on the *Quick Inventory of Depressive Symptomatology Self-Report* ($QIDS\text{-}SR_{16}$). And only the patient without history of bipolar disorder or psychosis, with the first major depressive episode and stable medical conditions before 30-year-old were enrolled.[3] Other exclusion criteria include: failed to respond to any prior trial of an antidepressant currently; current pregnancy, not using contraception, or breastfeeding; unstable psychiatric or general medical conditions requiring hospitalization or contraindicate study medication; laboratory abnormalities; requirement of an anticonvulsant; treatment with electroconvulsive therapy, vagal nerve stimulation, repetitive transcranial magnetic stimulation or other somatic treatments in the current episode; current depression-specific psychotherapy, and significant suicide risk. Of the 309 subjects, 296 received at least one dose of either sertraline or placebo and underwent at least one baseline assessment (Table S1).

##### Section M2: Functional MRI acquisition and preprocessing

Resting-state functional magnetic resonance imaging (rs-fMRI) data were acquired by T2* weighted images using a single-shot gradient echo-planar pulse sequence. At each site, neuroimaging data were collected with 3 Tesla scanners lasting for 8 minutes with different scanners but the same parameter set (repetition time 2000 ms, echo time 28 msec, flip angle 90°, matrix size 64×64, resolution 3.2×3.2 $mm^2$, slice thickness 3.1 mm).

The acquired rs-fMRI data were preprocessed using the reproducible fMRIprep pipeline.[4] The T1 weighted image was corrected for intensity nonuniformity and then stripped skull. Spatial normalization was done through nonlinear registration, with the T1w reference.[5] Using FSL, brain tissue such as cerebrospinal fluid, white matter, and grey matter was segmented from the reference, brain-extracted T1 weighted image.[6] The fieldmap information was used to correct distortion in low-frequency and high-frequency components of fieldmap caused by field inhomogeneity. With less fieldmap distortion, a corrected echo-planar imaging reference was obtained from a more accurate co-registration with the anatomical reference. The BOLD reference was then transformed to the T1-weighted image with a boundary-based registration method, configured with nine degrees of freedom to account for distortion remaining in the BOLD reference.[7] Head-motion parameters (rotation and translation parameters of volume-to-reference transform matrices) were estimated with MCFLIRT (FSL). BOLD signals were slice-time corrected and resampled onto the participant's original space with head-motion parameters, susceptibility distortion correction, and then resampled into standard space, generating a preprocessed BOLD run in MNI152NLin2009cAsym space. Automatic removal of motion artifacts using independent component analysis (ICA-AROMA)[8] was performed on the preprocessed BOLD time-series in MNI space after removal of non-steady-state volumes and spatial smoothing with an isotropic Gaussian kernel of 6 mm FWHM (full-width half-maximum).

Then, we reviewed the imaging parameters of each participant. With quality control for head motion (>0.5 mm motion framewise displacement or BOLD signal displacements > 0.5%)[9, 10], finally, the rs-fMRI data from 131 patients with sertraline treatment and 144 patients with placebo treatment were usable. Patients in these two arms showed no significant differences in the sociodemographic and clinical variables (Figure S4).

##### Section M3: Clinical measurements

The clinical outcome was estimated using the *Hamilton Depression Rating Scale* ($HAMD_{17}$). Other clinical measurements used in our study included: total score, general distress subscale score (gd), anhedonic depression subscale score (ad) and anxious arousal subscale score (aa) of *Mood and Anxiety Symptom Questionnaire* (MASQ); neuroticism score (ne), extraversion score (ex), openness score (op), agreeableness score (ag), conscientiousness score (co) in *NEO-Five Factor Inventory* (NEO); total score of *Mood Disorders Questionnaire* (MDQ); total score of *Quick Inventory of Depressive Symptomatology*

*Self Report* (QIDS). Total score of pre-treatment (pre) and post-treatment (post) *State–Trait Anxiety Inventory* (STAI); *Concise Health Risk Tracking Propensity score* (CHRTP); total score of *Self-Administered Comorbidity Questionnaire* (SCQ). Total score of *Snaith-Hamilton pleasure scale* (SHAPS); total score of *Social Adjustment Scale* (SAS); total score of *Standardized Assessment of Personality Abbreviated Scale* (SAPAS); total score of *Anger Attacks Questionnaire* (AAQ); total score, emotional abuse subscale score (ea), emotional neglect subscale score (en), physical abuse subscale score (pa), physical neglect subscale (pn), and sexual abuse subscale score *Childhood Trauma Questionnaire* (CTQ). For participants lacking an endpoint $HAMD_{17}$, the $HAMD_{17}$ at baseline, at week 1, at week 2, at week 3, at week 4, at week 6, baseline QIDS total score, baseline MASQ, Anhedonic Depression and General Distress, SHAPS, age, years of education, sex and Wechsler Abbreviated Scale of Intelligence t-scores for Vocabulary and Matrix Reasoning were used to fit Bayesian regression to impute endpoint $HAMD_{17}$ value.[2]

#### Section M4: Common orthogonal basis extraction (COBE)

COBE is a well-established algorithm based on group component analysis to extract the common basis for multi-block data.[11] COBE has been successfully applied to extract dissimilarity-maximized feature patterns from rs-fMRI for defining heterogeneous subtypes in healthy populations and prediction of cognitive and behavioral measures.[12, 13] Given the multi-block data $\boldsymbol{Y}_k$, it decomposes the data into common and individualized subspace by solving the optimization problem:

$$\min_{\overline{\boldsymbol{A}}^c,\breve{\boldsymbol{A}}_k^c} \sum_{c=1}^{C}\sum_{k=1}^{K} ||\boldsymbol{Y}_k - \overline{\boldsymbol{A}}^c\overline{\boldsymbol{Y}}_k^c - \breve{\boldsymbol{A}}_k^c\breve{\boldsymbol{Y}}_k^c||_F^2$$

$$s.t. \overline{\boldsymbol{A}}^{c^T}\overline{\boldsymbol{A}}^c = \boldsymbol{I}, \breve{\boldsymbol{A}}_k^{c^T}\breve{\boldsymbol{A}}_k^c = \boldsymbol{I}, \overline{\boldsymbol{A}}^{c^T}\breve{\boldsymbol{A}}_k^c = \boldsymbol{0}$$

where $\overline{\boldsymbol{A}}^c$ represent the loading corresponding to the *c*-th common component, $\overline{\boldsymbol{Y}}_k^c$ represent the corresponding coefficient matrix in *k*-th grouped data, and $\breve{\boldsymbol{A}}_k^c\breve{\boldsymbol{Y}}_k^c$ represent the projection of the *k*-th group of subjects in individual subspace. More specifically, in our research, FCs of whole data (*N* subjects) were randomly split into *K* groups. Each group had $N_k$ subjects, which was the maximized decomposition number. Then common feature $\overline{\boldsymbol{C}}$ across $K$ group data-blocks was extracted by COBE. Individualized FCs $\breve{\boldsymbol{F}}$ were acquired by removing $\overline{\boldsymbol{C}}$ from $\boldsymbol{Y}$. In our experiment, all patients who accepted different therapies were used together to gain $\overline{\boldsymbol{C}}$ from COBE decomposition since the algorithm is unsupervised and the t-test results showed that all MDDs with different treatments had no significantly different FCs. The hyperparameter $K$ was set to 10 and the number of COBE decomposed components ($C$) was set to 5. One reason was that the hyperparameter $K$ made a negligible influence on the decomposition patterns (Figure S2). When hyperparameter was set to 5, 10, and 20, all of them had relatively highly stable decomposed results before the 6-th component. The 5th component pattern visualizations with grouped hyperparameters 5, 20 were shown in Figure S2 (B, D), with grouped hyperparameter 10 were shown in Figure S7 E. All the common components projected matrices computed from differently grouped hyperparameters were very similar. Their Pearson's correlation coefficients ($r$) were larger than 0.9. Other COBE-like Joint and Individual Variation Explained methods were applied in joint matrix decomposition for $K$ data blocks and in individual matrix decomposition for $N_k$ samples in each block simultaneously.[11, 14] In those methods, a too large $K$ limited the maximum numbers of decomposition, which is assumed to extract inadequate common features. A too small $K$ might weaken the ability to extract common features via join matrix decomposition. Therefore, following the $K$ setting in other multi-block data analyses,[15, 16] we empirically set $K = 10$ finally to keep a balance in the size of $K$ and $N_k$. Another hyperparameter, the number of COBE decomposed components ($C$), was set to 5. The sixth decomposed common component had a significant correlation-ship to clinical treatment (Figure S5E, F). So, it might explain why removing the sixth or more common components, the prediction performance dramatically decreased for placebo and sertraline treatment outcome change prediction (Figure S5B, C). Another reason was that the first five decomposed patterns were relatively stable, explaining near 70% common covariance (Figure S5A, D).

#### Section M5: Sparse connectome predictive modeling

With individualized FCs, we will perform sparse connectome predictive modeling for treatment outcome prediction. The prediction model combines connectome-based predictive modeling (CPM)[17] and LASSO regression. CPM was a recently developed method for identifying informative brain connectivity in behavior prediction tasks.[17] Neuroimaging studies have shown its strength in detecting predictive connectivity features for individual creative ability and intelligence.[18, 19] In the CPM step, Pearson's correlation coefficient between clinical outcome (measured as pre- minus post-treatment $HAMD_{17}$ score) with each connectivity in the brain connectome matrix across participants was calculated. The edges showing significant correlation (below a threshold $\lambda$) to the clinical outcome were then retained for the subsequent prediction analysis. To further derive a compact feature pattern that is less sensitive to overfitting and more interpretable, we trained a sparse linear regression model with L1 regularization (LASSO) based on the CPM-informed FC features. The prediction performance was evaluated by 10×10-fold cross-validations (CVs). Specifically, for each 10-fold CVs, the data was randomly partitioned into ten folds. One fold was left out as the test data, while the remaining nine folds were used as the training data. For those subjects who have rs-fMRI scans of two runs, both scans were assigned to either training set or test set. The prediction results of two runs were then averaged as the final prediction of this subject. The process was then repeated ten times, where each of the ten folds was used exactly once as test data to obtain predicted $HAMD_{17}$ score changes for all subjects. To enhance the stability of the prediction, the above 10-fold process was repeated 10 times. From 10 repetitions, the median of the resulting predicted $HAMD_{17}$ score changes of each participant was used as the final prediction. Furthermore, the specificity of the model was tested by using models trained from 100 folds to predict the outcome in the other treatment arm. The optimal values of the CPM parameter $\lambda$ ([0.005, 0.01, 0.05, 0.1]) and L1 regularization parameter $\alpha$ ([1, 0.5, 0.1, …, 0.001]) were determined using an inner-loop 10-fold cross-validation on the training set (Figure S12).[20] R-squared value ($R^2$) and Pearson's correlation coefficient were calculated between the predicted outcomes and the observed ones as performance metrics to evaluate the model efficacy. For each treatment predictive model, with cross-treatment prediction experiments, eight correlation coefficients between true treatment response and predicted treatment response training from individualized FC and raw FC were false discovery rate (FDR) corrected.

#### Section M6: Effect analysis of functional connectivity individualization

To examine the effects of FC individualization on improving treatment outcome prediction, we applied the Wilcoxon signed-rank test to compare $R^2$ values and Pearson correlation coefficients of ten repeated runs of 10-fold CVs of individualized FCs and raw FCs. The statistical significance in performance comparison was corrected using FDR. We further investigated why individualized FCs enhanced the prediction performance from feature and model perspectives, respectively. First, from a model perspective, we used the Wilcoxon signed-rank test to detect the significantly changed weights in the Lasso model trained using individualized FCs versus raw FCs, across 100 folds. The model weights trained from individualized FCs and raw FCs were on the same scale, so they could be compared directly. FDR was used to correct the significance of the change in prediction weight for each FC. Then, from a feature perspective, the change of Pearson's correlation coefficient between the $HAMD_{17}$ change and predictive FCs was evaluated by the Wilcoxon signed-rank test in the CPM step. For each pairwise statistical comparison, the correlation coefficients of individualized FCs and raw FCs across 100 folds were utilized. Then FDR was used to correct the significance of the change in correlation between each FC and $HAMD_{17}$ change. Finally, after FC individualization, the inconsistent connectivity pattern was defined as the pairwise FCs with significantly changed prediction weight as well as with significantly changed correlation coefficient to $HAMD_{17}$ change.

#### Supplementary results

**Figure S1. Illustration of COBE algorithm.** The FCs of all subjects were randomly split into $K$ groups first. Then COBE extracted total $C$ common feature $\bar{\boldsymbol{C}}$. Individualized FC ($\breve{\boldsymbol{F}}$) was obtained by subtracting all common feature $\bar{\boldsymbol{C}}$ from raw FC ($\boldsymbol{Y}$).

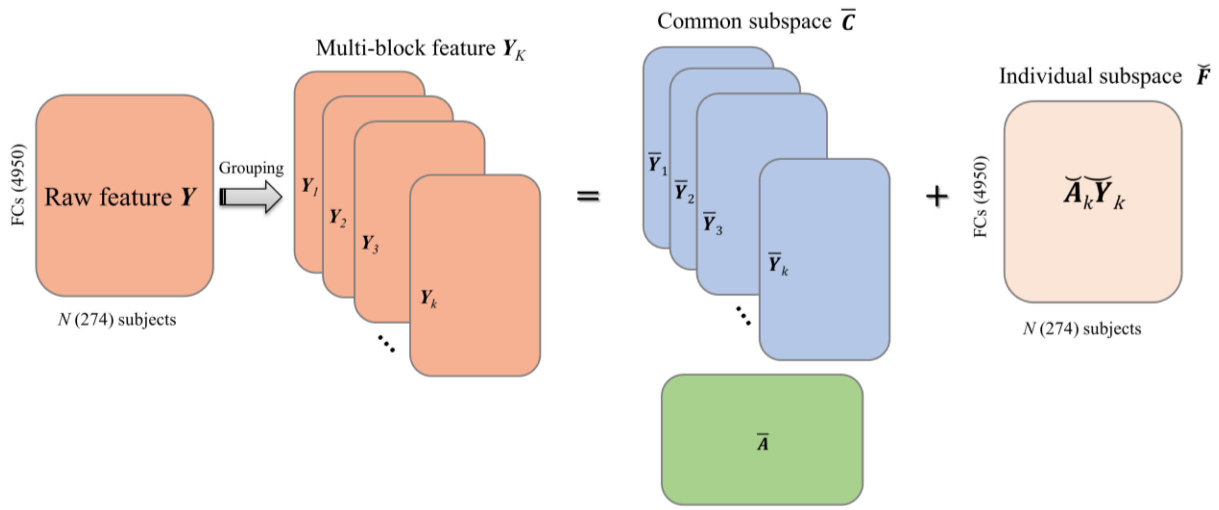

**Figure S2. The COBE decomposition pattern using different group splitting hyperparameter** $K$. When the group parameter ($K$) was set to 5, **A** the stability analysis of COBE decomposition pattern was evaluated by calculating the correlation coefficient of COBE transformed weights across 100 folds, and **B** the heatmap of the 5th component transformation average weights in 100 folds was shown. When the group parameter was set to 20, **C** the stability analysis of COBE decomposition pattern was evaluated by calculating the correlation coefficient of the COBE transformed weights across 100 folds. and **D** the heatmap of the 5th component transformation average weights in 100 folds was shown.

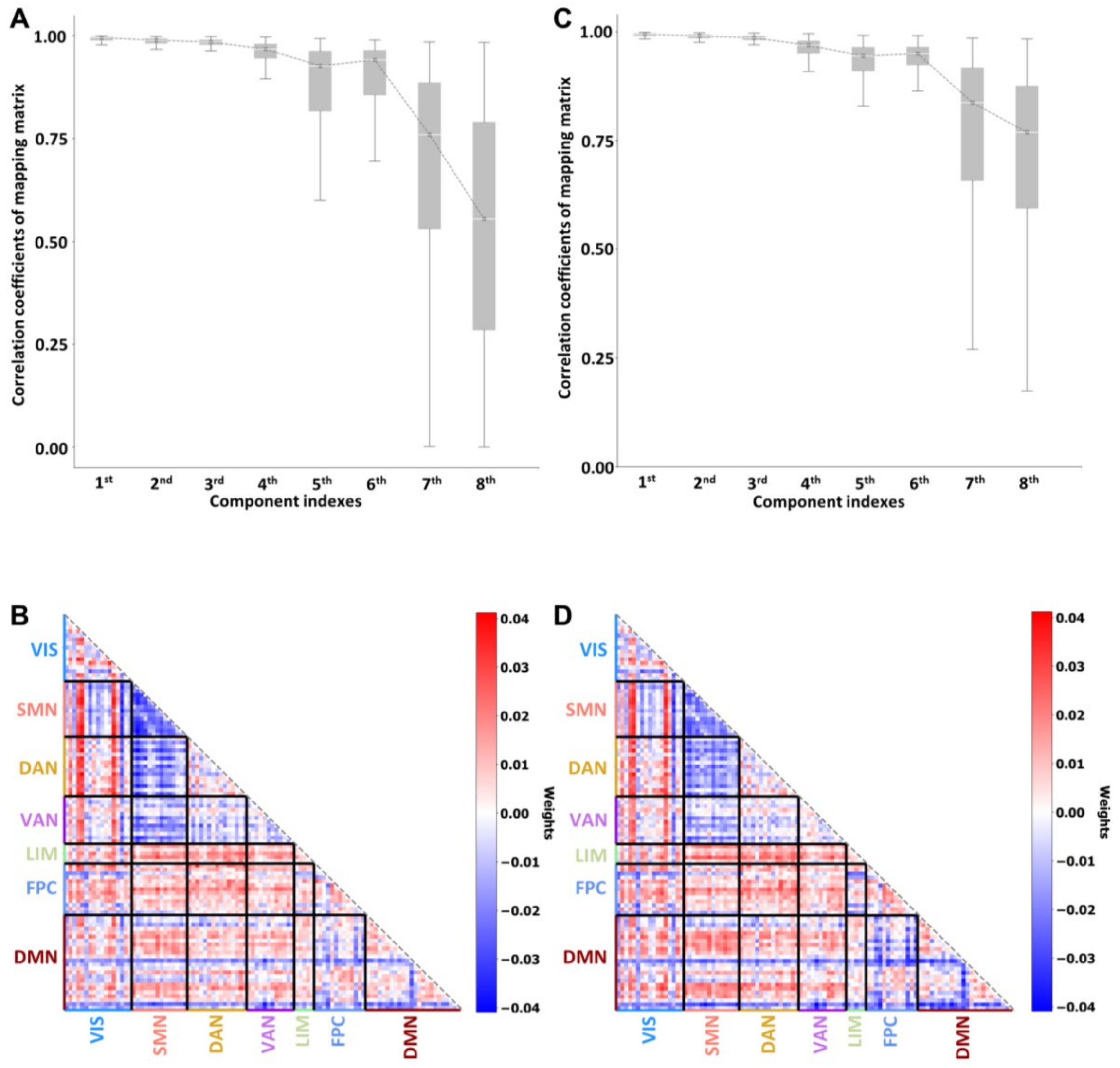

**Figure S3. Framework of our study. A** Regional time series were extracted from fMRI BOLD signals based on the Schaefer atlas. **B** FCs were obtained by computing the Pearson's correlation coefficient of time series between any pair of ROIs. They were then individualized by removing common components using the COBE decomposition. The raw FC-based model was trained with non-individualized FC features, from step **B**① to step **C**② directly. **C** CPM feature selection. Only the FCs significantly correlated to $HAMD_{17}$ change were kept for the subsequent prediction analysis. **D** Lasso regression-based predictive modeling for treatment outcome prediction and biomarker quantification.

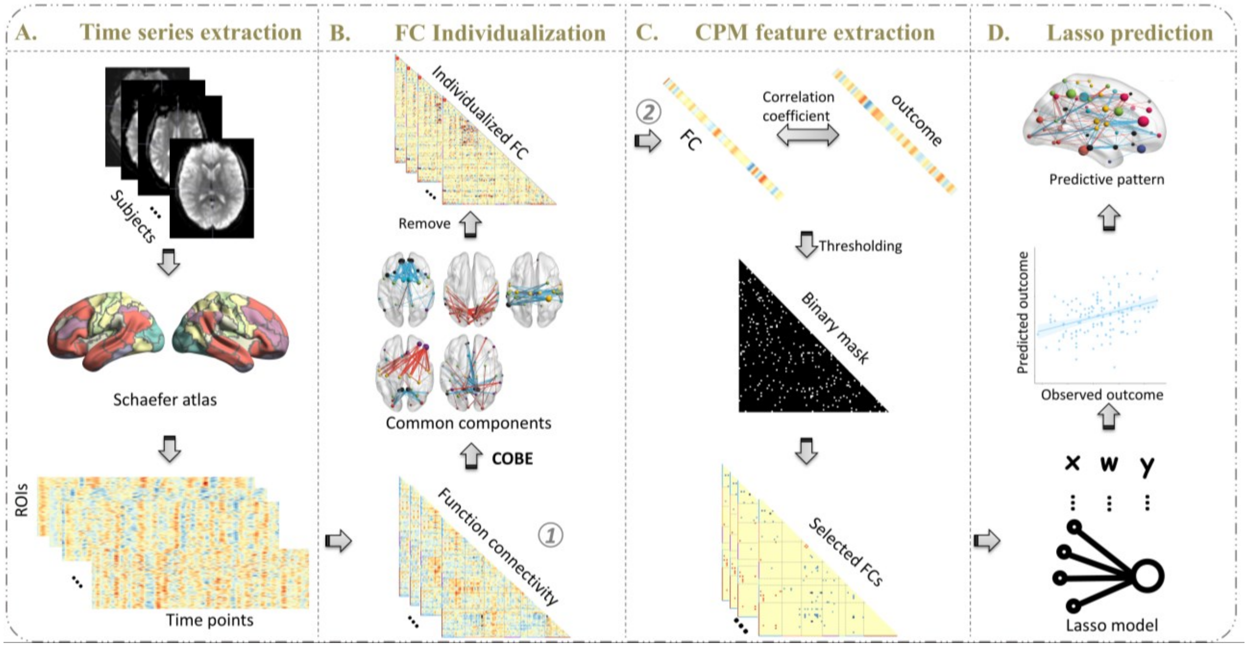

**Figure S4. Patient demographics.** The patients received different treatments had no significant difference in demographics, including site, sex, education, age, baseline $HAMD_{17}$ score and race. All *p* values were corrected by FDR. Red color was related to the patients with placebo treatment and blue color was related to the patients with sertraline treatment.

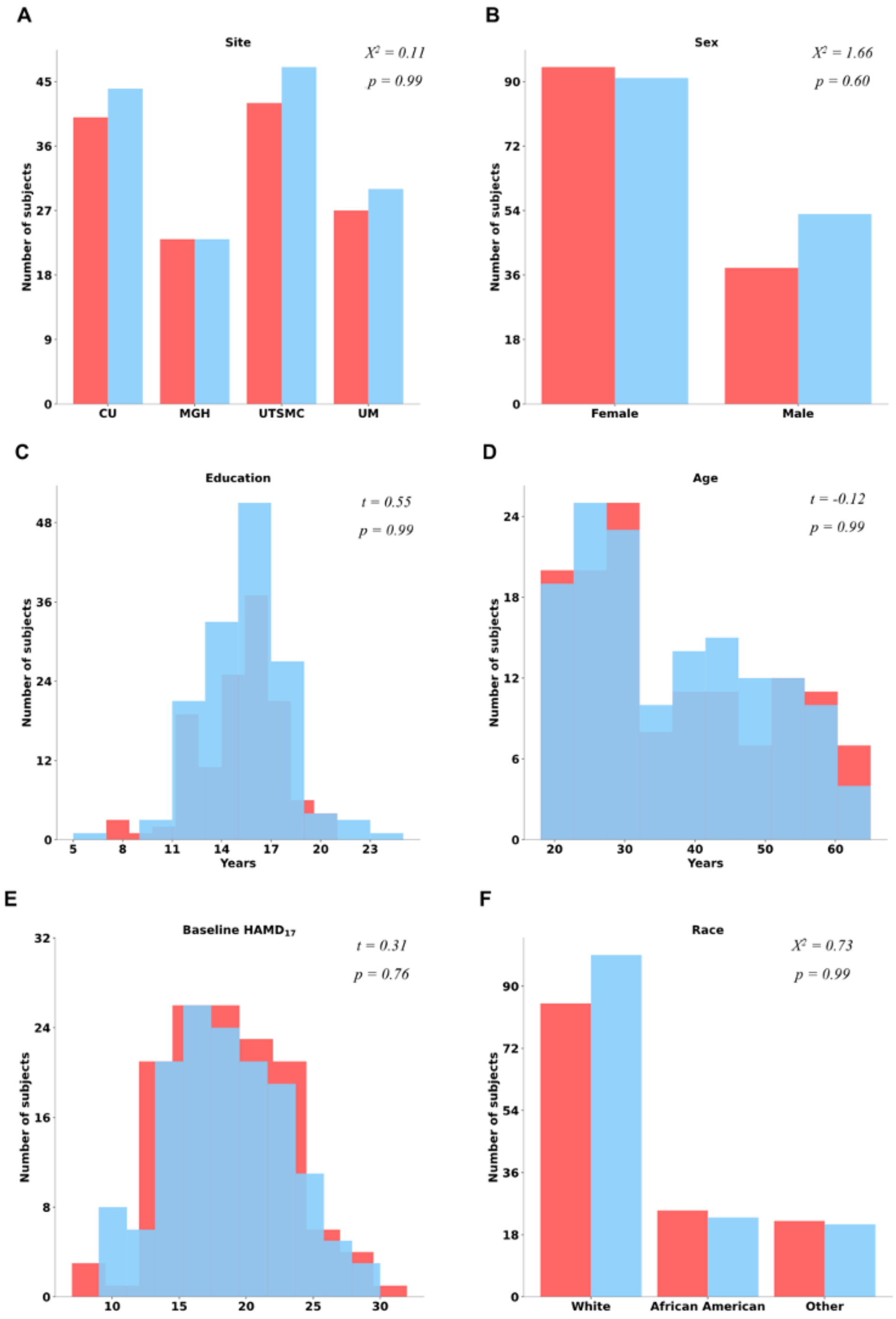

**Figure S5. Effects of varying the number of common FC components to extract and exclude when *K* = 10. A** The covariance explained ratio plot of common components. The first five components were circled and the dasher line indicated that these five components explained more than 69% accumulated covariance. **B, C** $R^2$ between the prediction treatment outcome training with FC individualized from different numbers of the COBE decomposition and the true treatment outcome. **B)** placebo arm, and **C)** sertraline arm. **D** Stability analysis of COBE decomposition pattern was evaluated by calculating the correlation coefficient of COBE transformed weights across 100 folds. The first six components had a median correlation coefficient larger than 0.9. **E, F** Pearson correlation coefficient between each common feature transformed from COBE decomposition and $HAMD_{17}$ score change with different therapies (significance of correlation: ⋆$p$<=0.05, ⋆⋆$p$<=0.01, ⋆⋆⋆$p$<=0.001). **E)** placebo arm, **F)** sertraline arm.

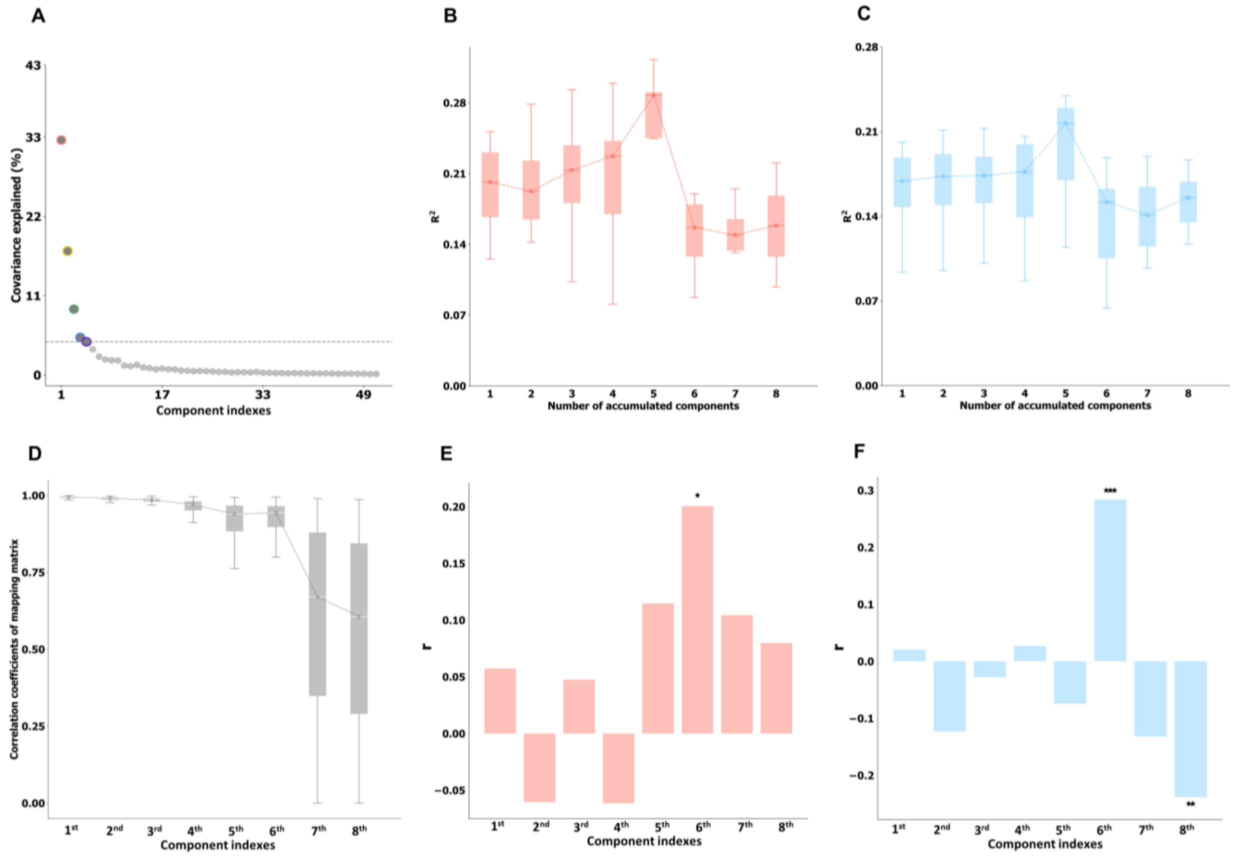

**Figure S6. COBE decomposition mapping weight of the first five common components.** **A-E** Connectivity patterns of the first five common components, obtained by averaging the mapping weights across all runs of cross-validation. They were sorted from **A** to **E** ascendingly (**A** showed the first component). Only the top 40 connections were visualized for the interpretation of each common component. Blue color: negative weight. Red color: positive weight. Thicker link. larger weight absolute value. VIS, visual network; SMN, somatomotor network; DAN, dorsal attention network; VAN, ventral attention network; LIM, limbic network; FPC, frontoparietal control network; DMN, default mode network.

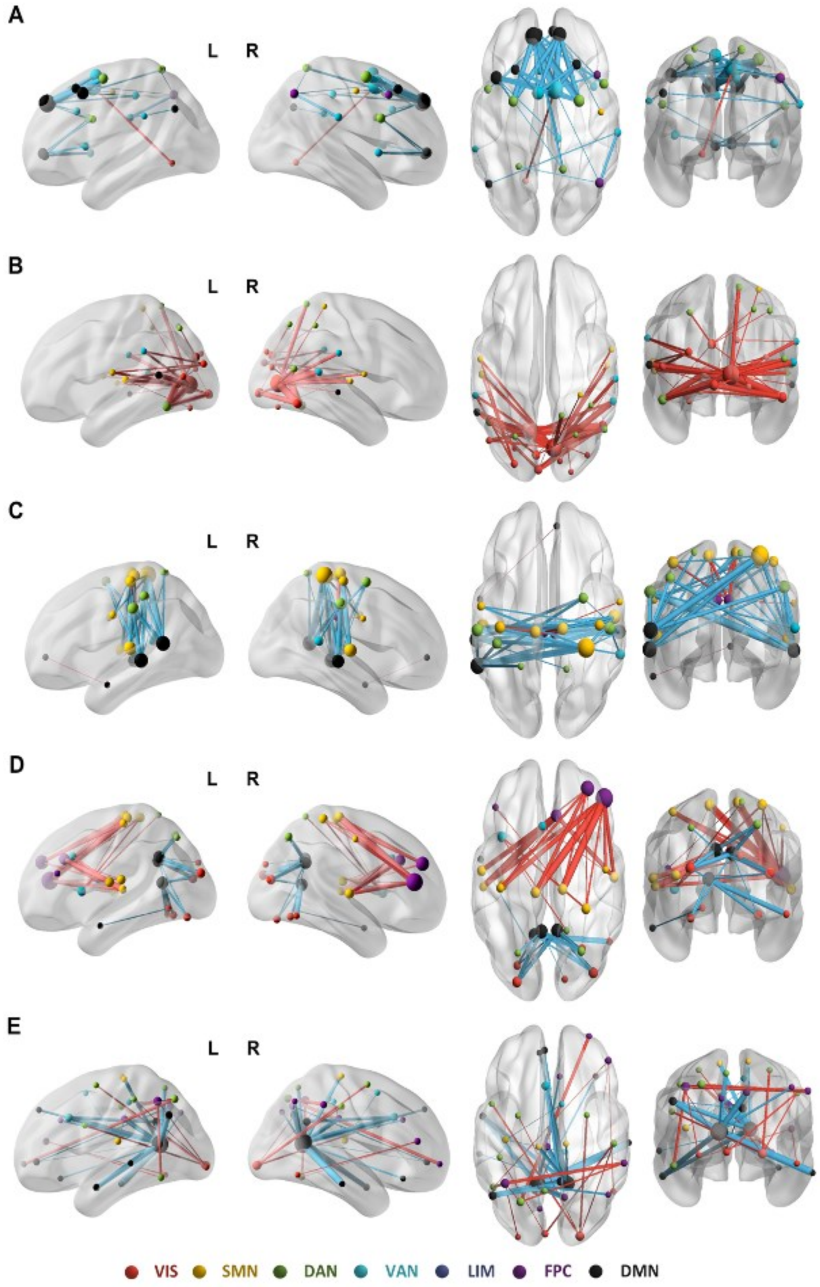

**Figure S7. Heatmap of COBE decomposition mapping weights of the first five composition.** The panels order from **A** to **E** were same with the panels in Figure S6. **A** was the first component, **E** was the fifth component.

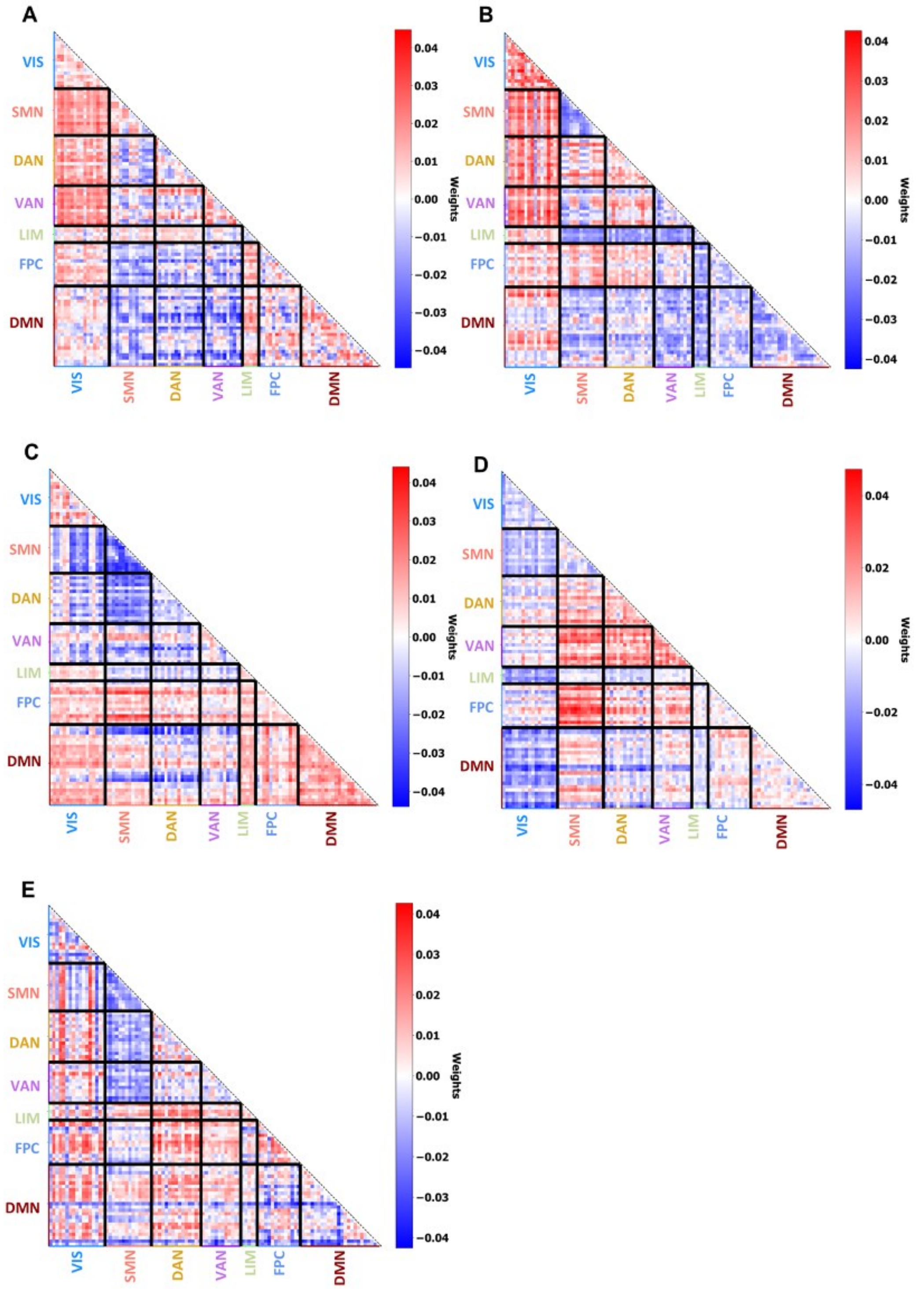

**Figure S8. The reconstructed FCs from the first 5 COBE common components.** On the top panel, the top 40 connections of the mean of the reconstructed FCs for all patients were plotted. Deeper red and thicker link corresponded to higher averaged FCs. On the bottom, all reconstructed FCs were summed as node strength rendered on the cortical surface. Deeper red regions corresponded to higher node strength.

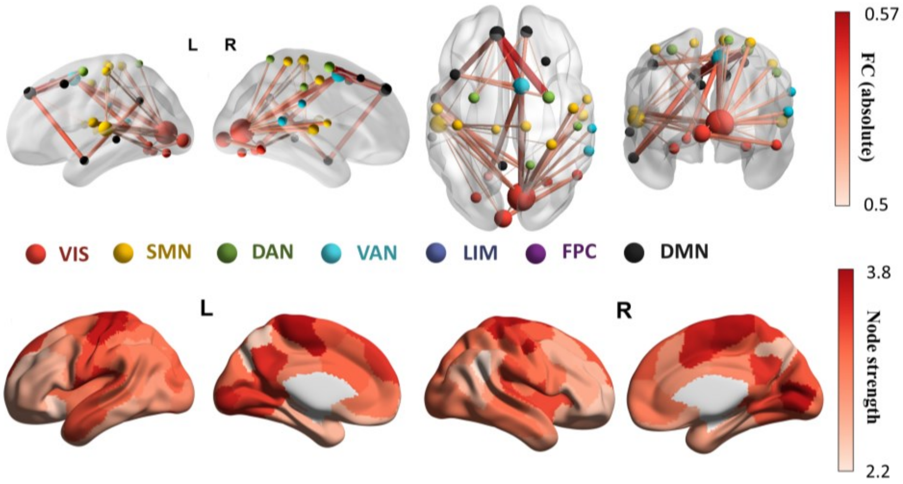

**Figure S9. Cross-treatment arm prediction of treatment outcome.** Models were trained with **A** individualized FCs and **B** raw FCs from the sertraline treatment group and then tested on the placebo group. Models were trained with **C** individualized FCs and **D** raw FCs from the placebo treatment group and then tested on the sertraline group. No significant prediction performance was found for all these conditions.

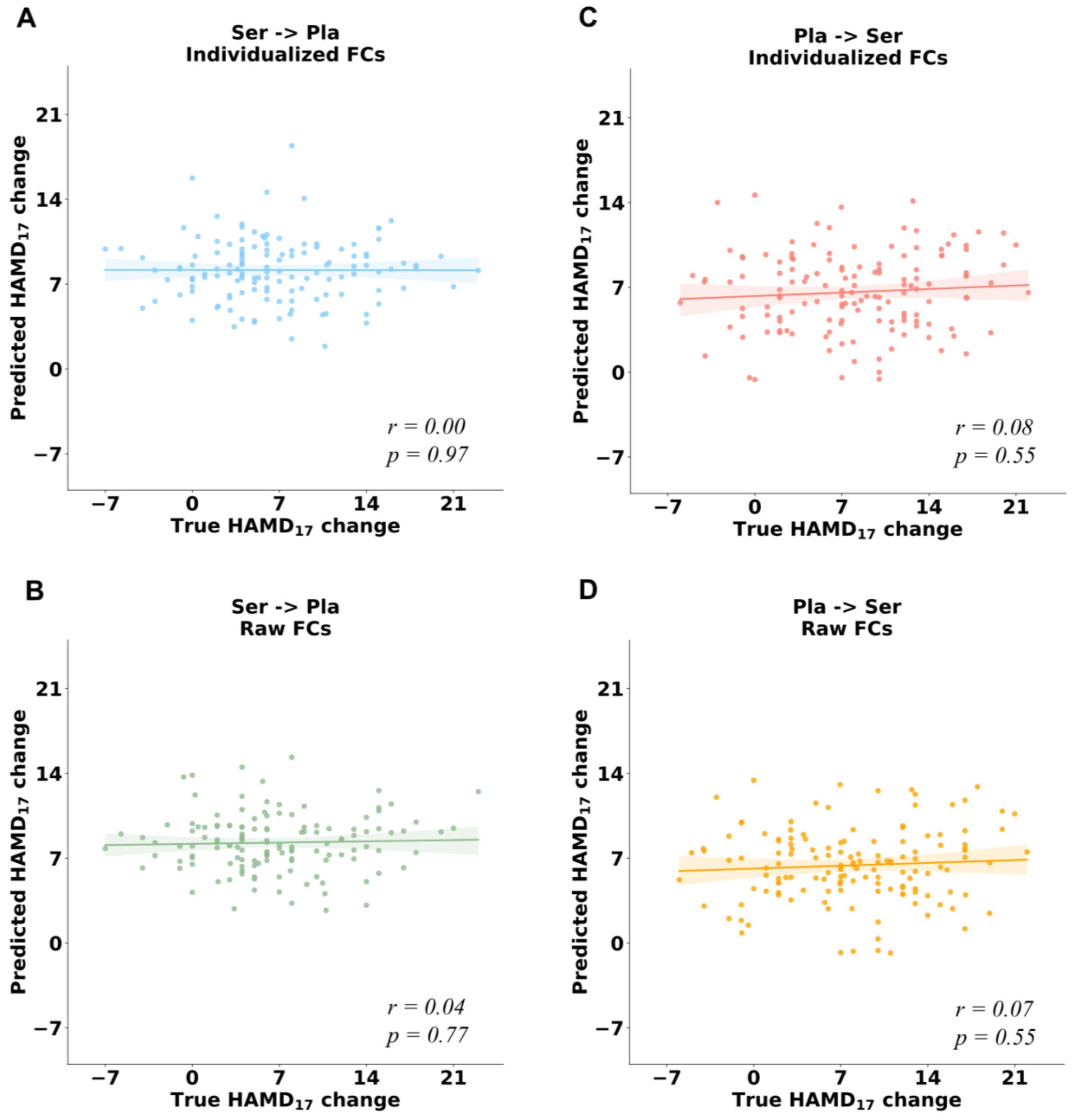

**Figure S10. Prediction of the outcome specific to sertraline and placebo arm in 10 × 10 cross-validations using common FCs reconstructed from common components.** The p values were computed from permutation test.

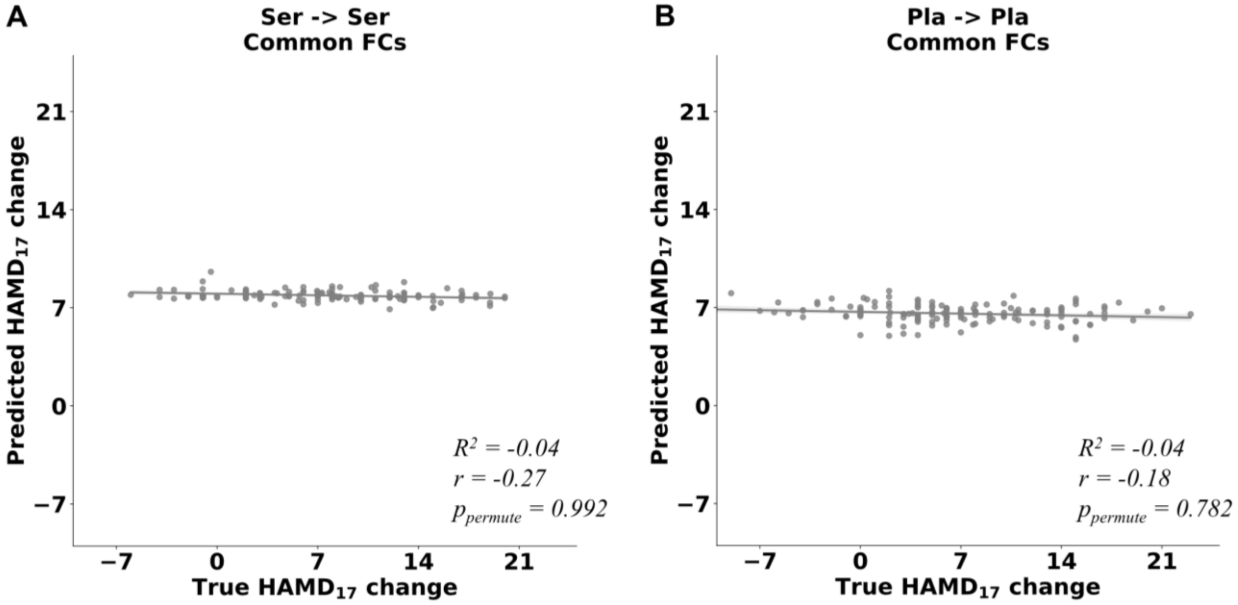

**Figure S11. Two examples to illustrate the time-by-FC effects in sertraline arm.** They were examples of the visualization of Table S4. The *p* values were FDR corrected.

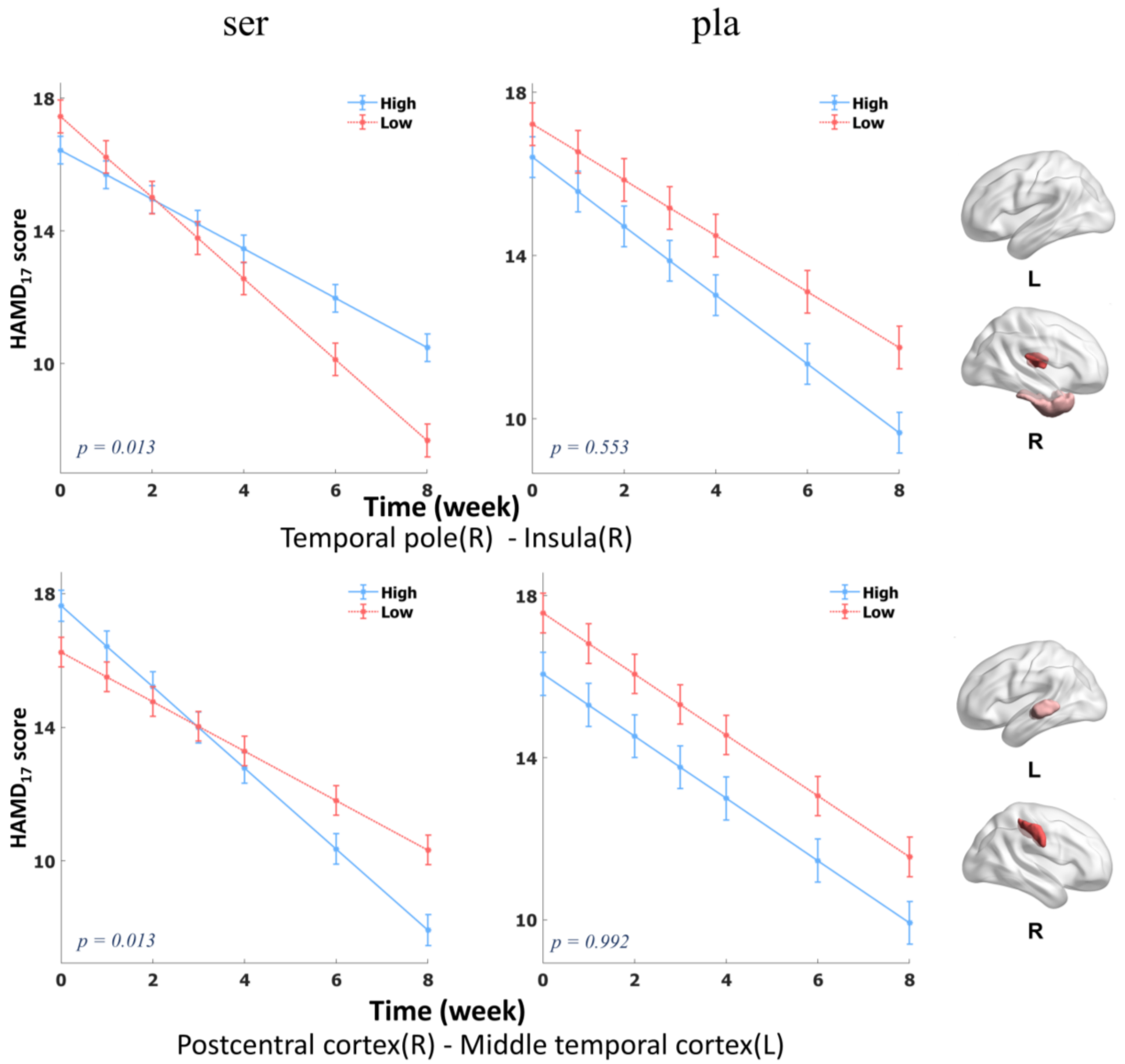

**Figure S12. Two examples to illustrate the time-by-FC effects in placebo arm.** They were examples of the visualization of Table S5. The *p* values were FDR corrected.

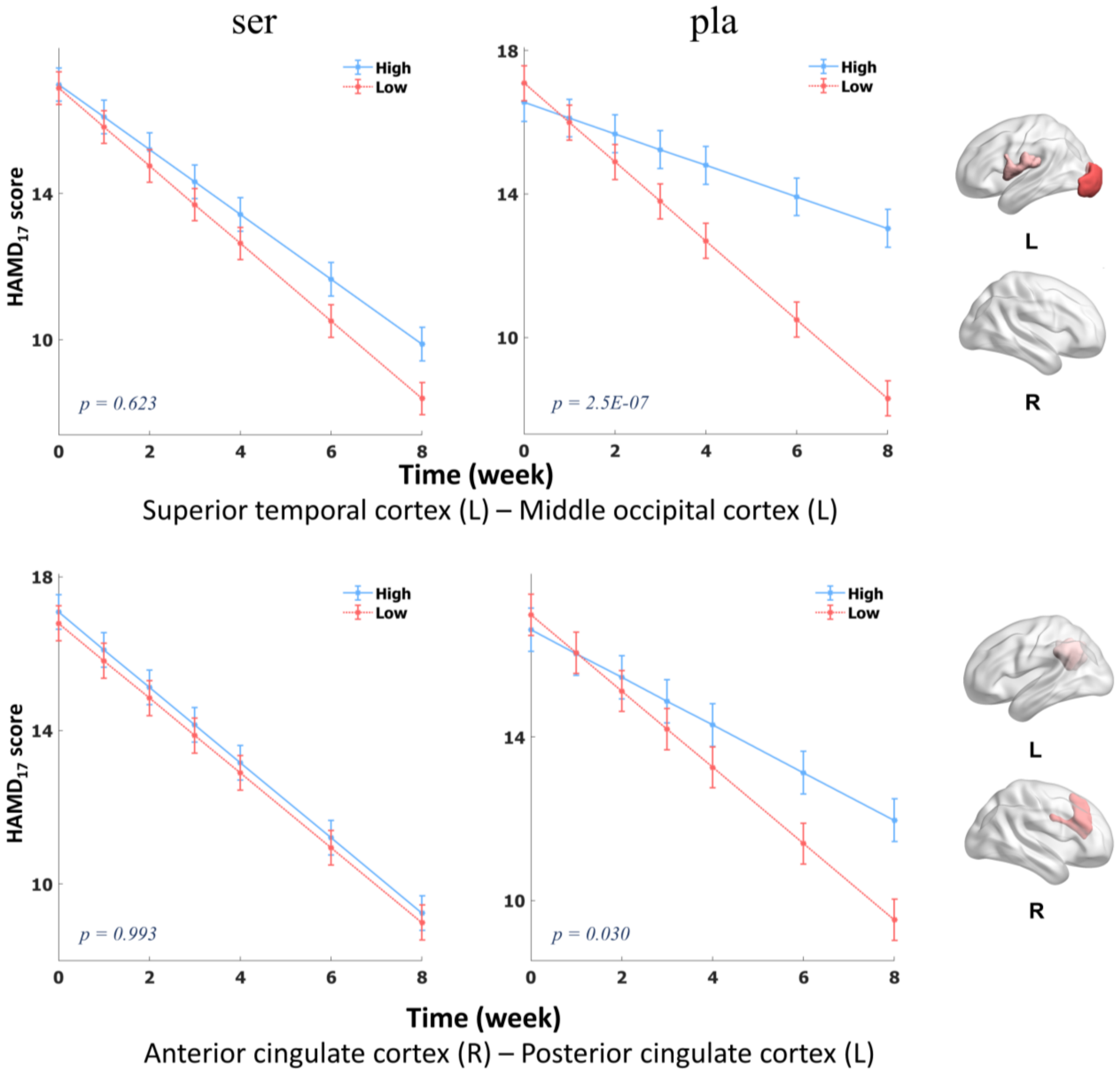

**Figure S13. The model performance of different hyperparameter settings in the CPM feature selection step and the Lasso regression model.** *r* values between predicted score and true score were used as evaluation metric. As shown in panel **A**, when $\lambda = 0.005$ and $\alpha = 0.1$, the sertraline treatment predictive model had the best performance. And in panel **B** when $\lambda = 0.05$ and $\alpha = 0.5$, the model had the best prediction performance to placebo treatment.

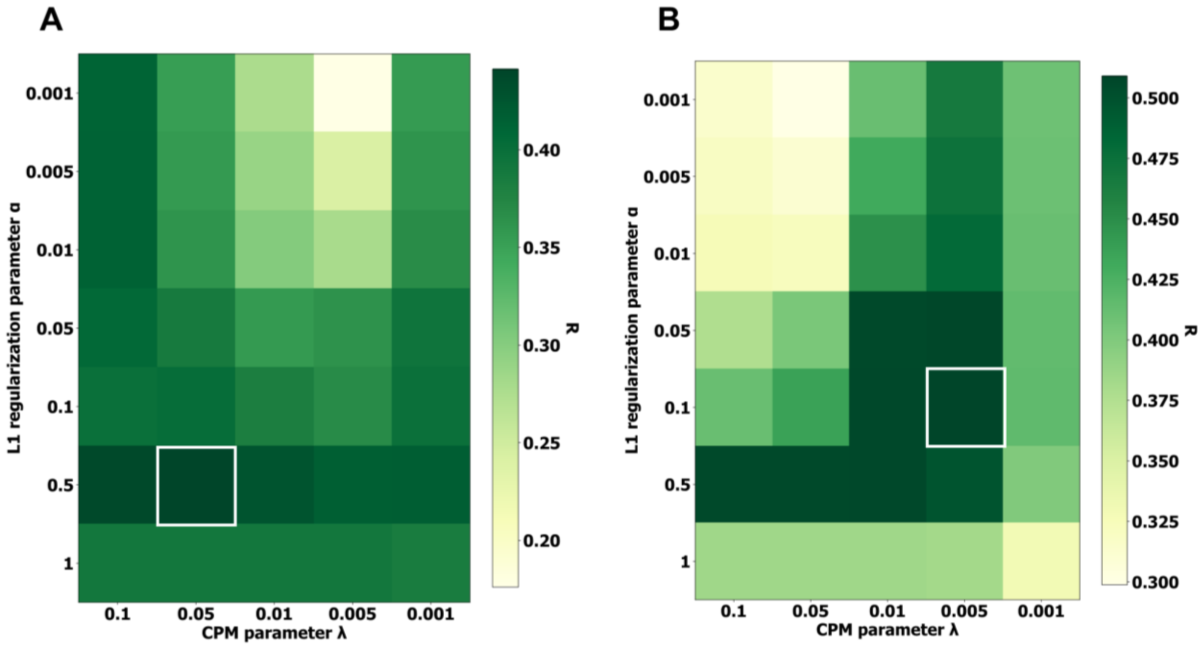

**Figure S14. Treatment outcome predictions in leave-study-site-out cross validation.** CU, Columbia University; MG, Massachusetts General Hospital; TX, University of Texas Southwestern Medical Center; UM, University of Michigan. **A** The leave-one-site CV for sertraline treatment with Combat correction, $R^2 = 0.11$, Pearson's $r = 0.42$, FDR corrected $p = 8.0 \times 10^{-7}$ based on the one-sided test against the alternative hypothesis that $r > 0$. **B** The leave-one-site CV for placebo treatment with Combat correction, $R^2 = 0.10$, Pearson's $r = 0.36$, FDR corrected $p = 8.6 \times 10^{-6}$ based on the one-sided test against the alternative hypothesis that $r > 0$. **C** The leave-one-site CV for placebo treatment with z-score normalization, $R^2 = 0.10$, Pearson's $r = 0.33$, FDR corrected $p = 1.1 \times 10^{-4}$ based on the one-sided test against the alternative hypothesis that $r > 0$. **D** The leave-one-site CV for placebo treatment with z-score normalization, $R^2 = 0.09$, Pearson's $r = 0.31$, FDR corrected $p = 1.3 \times 10^{-4}$ based on the one-sided test against the alternative hypothesis that $r > 0$.

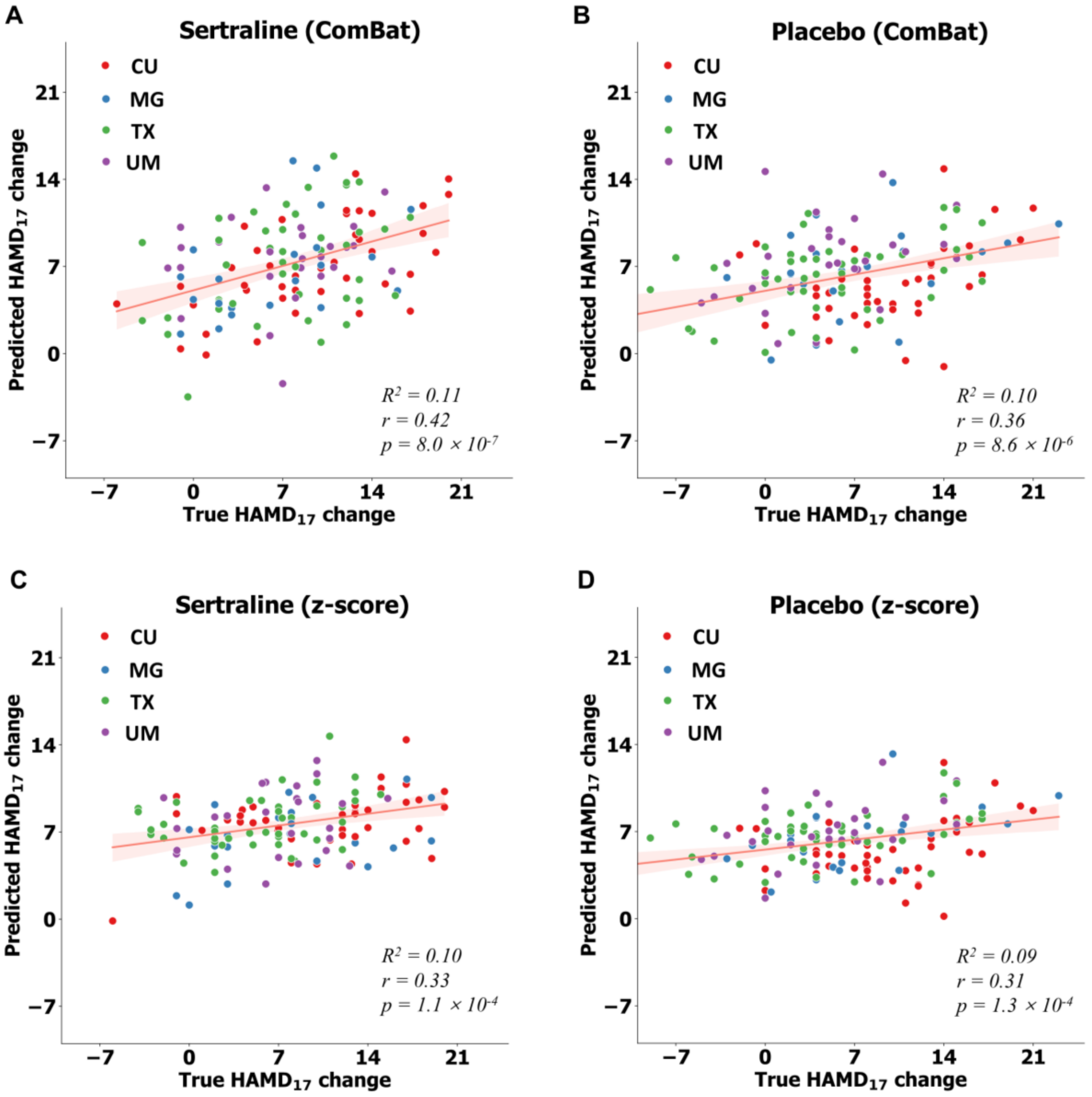

**Figure S15. Treatment outcome predictions in 10 × 10 cross validation, training with the clinical measurements and demographic information.** **A** Prediction for placebo treatment, Pearson's $r = 0.13$, FDR corrected $p = 0.14$. **B** Prediction for sertraline treatment, Pearson's $r = 0.08$, FDR corrected $p = 0.38$.

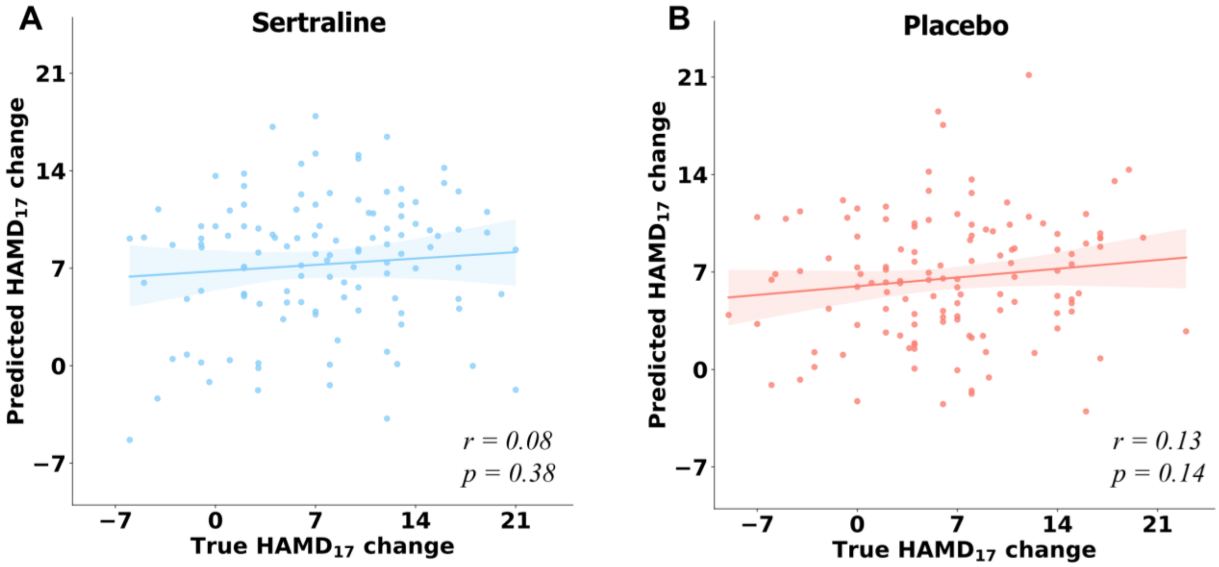

**Figure S16. The difference of prediction performances training from raw FCs and individualized FCs.** It was evaluated by Wilcoxon signed-rank test. *p* values were corrected from FDR. **A** Difference of $R^2$ in sertraline treatment ($w_{individualized\ vs\ raw} = 2.57$, $p_{individualized\ vs\ raw} = 0.014$; $w_{raw\ vs\ common} = 3.78$, $p_{raw\ vs\ common} = 0.001$). $R^2_{mean}$ and $R^2_{std}$ for raw FCs prediction model were 0.15 and 0.03. $R^2_{mean}$ and $R^2_{std}$ for individualized FCs prediction model were 0.20 and 0.04. $R^2_{mean}$ and $R^2_{std}$ for common FCs prediction model were -0.03 and 0.007. **B** Difference of $R^2$ in placebo treatment ($w_{individualized\ vs\ raw} = 3.02$, $p_{individualized\ vs\ raw} = 0.006$; $w_{raw\ vs\ common} = 3.78$, $p_{raw\ vs\ common} = 0.001$). $R^2_{mean}$ and $R^2_{std}$ for raw FCs prediction model were 0.17 and 0.02. $R^2_{mean}$ and $R^2_{std}$ for individualized FCs prediction model were 0.26 and 0.05, $R^2_{mean}$ and $R^2_{std}$ for common FCs prediction model were -0.02 and 0.01. **C** Difference of *r* in sertraline treatment ($w_{individualized\ vs\ raw} = 2.72$, $p_{individualized\ vs\ raw} = 0.014$; $w_{raw\ vs\ common} = 3.78$, $p_{raw\ vs\ common} = 0.001$). $r_{mean}$ and $r_{std}$ for raw FCs prediction model were 0.38 and 0.03. $r_{mean}$ and $r_{std}$ for individualized FCs prediction model were 0.44 and 0.04, $r_{mean}$ and $r_{std}$ for common FCs prediction model were -0.201 and 0.04. **D** Difference of *r* in placebo treatment ($w_{individualized\ vs\ raw} = 3.18$, $p_{individualized\ vs\ raw} = 0.004$; $w_{raw\ vs\ common} = 3.78$, $p_{raw\ vs\ common} = 0.001$). $r_{mean}$ and $r_{std}$ for raw FCs prediction model were 0.42 and 0.02. $r_{mean}$ and $r_{std}$ for individualized FCs prediction model were 0.51 and 0.05, $r_{mean}$ and $r_{std}$ for common FCs prediction model were -0.199 and 0.07. The $r_{mean}$ and $R^2_{mean}$ here were slightly smaller than *r* and $R^2$ in the scatter plot (Figures 1 and 2). All the *p* values computed from eight comparison shown in four panels were corrected by FDR. For the prediction performance in scatterplot, in order to visualize the predictive $HAMD_{17}$ change clearly, we first calculated the median of predictive $HAMD_{17}$ changes in 10 × 10-fold CVs. However, in histogram plot, to verify that the improvement of model prediction was generalized across different randomized settings, we calculated the $R^2$ and *r* from test set in each 10-fold cross validation firstly. Then *r* and $R^2$ in 10 trials would be compared. The higher results in scatterplot were from the ensemble learning effect.[21, 22] Ensemble learning effect is an observation that fusing the prediction results from several weak machine learning models leads to better performance. For scatter plot, it integrated the results from all models in 10 × 10-fold CVs, thus, higher $r$ and $R^2$ were expected.

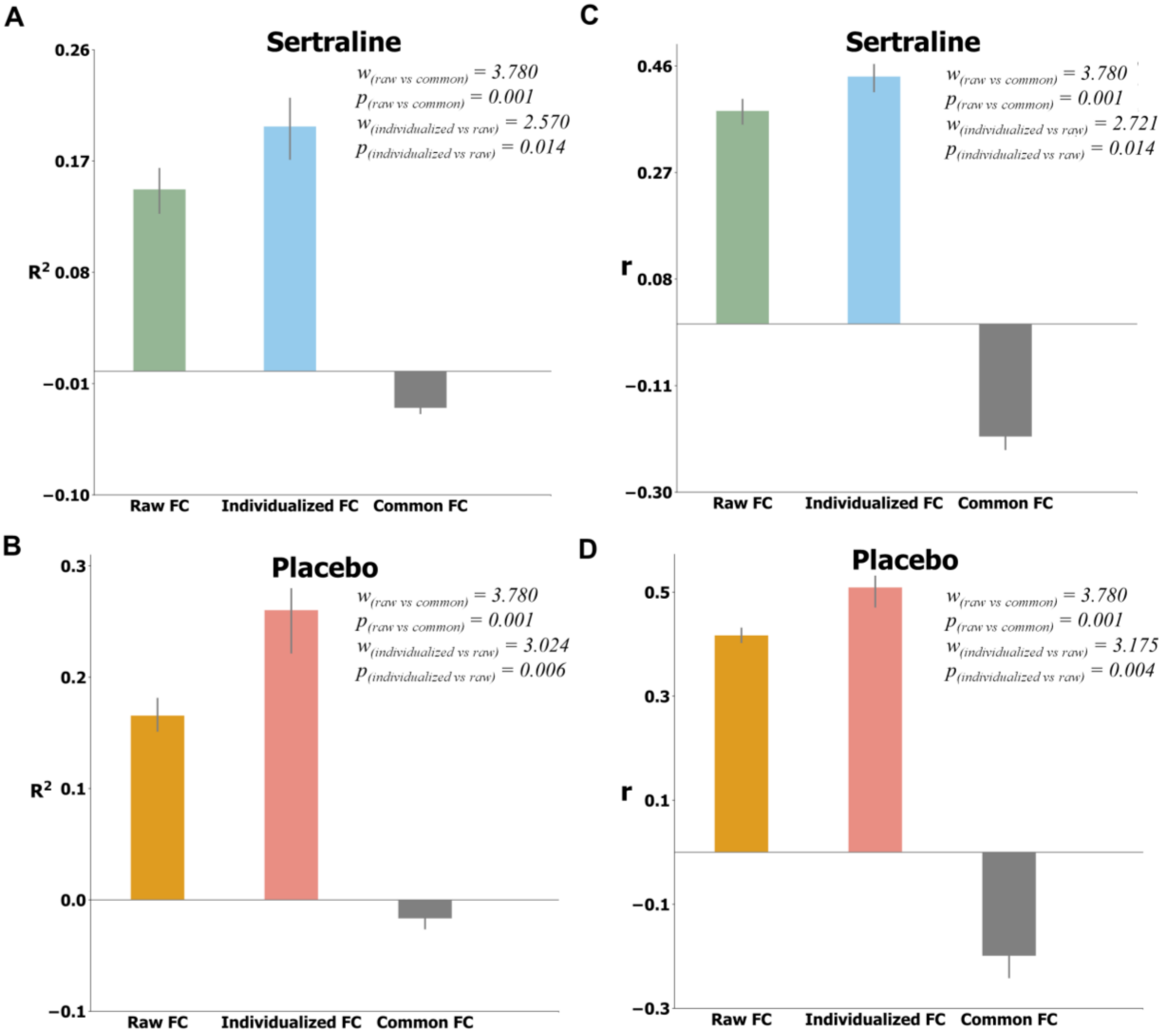

**Table S1. A summary of the correlation of transformed features from each COBE component and some clinical measurements.** Pearson correlation between each behavior score and each common component transformed feature was computed. *Mood and Anxiety Symptom Questionnaire* (MASQ); *NEO-Five Factor Inventory* (NEO); *Mood Disorders Questionnaire* (MDQ); *Quick Inventory of Depressive Symptomatology Self Report* (QIDS). *State–Trait Anxiety Inventory* (STAI); *Concise Health Risk Tracking Propensity score* (CHRTP); *Self-Administered Comorbidity Questionnaire* (SCQ). *Snaith-Hamilton pleasure scale* (SHAPS); *Social Adjustment Scale* (SAS); *Standardized Assessment of Personality Abbreviated Scale* (SAPAS); *Anger Attacks Questionnaire* (AAQ); *Childhood Trauma Questionnaire* (CTQ). The significant correlation coefficients ($p<0.05$ before FDR correction) were bolded.

| **Component index** | **1st** | | **2nd** | | **3rd** | | **4th** | | **5th** | |
|---|---|---|---|---|---|---|---|---|---|---|
| | ***r*** | ***p*** | ***r*** | ***p*** | ***r*** | ***p*** | ***r*** | ***p*** | ***r*** | ***p*** |
| MASQ (gd) | **0.150** | **0.012** | -0.046 | 0.444 | -0.001 | 0.981 | 0.077 | 0.197 | 0.029 | 0.627 |
| MASQ (ad) | 0.074 | 0.217 | -0.101 | 0.091 | **0.162** | **0.006** | 0.005 | 0.938 | 0.000 | 0.996 |
| MASQ (aa) | -0.013 | 0.823 | **0.131** | **0.028** | 0.015 | 0.797 | 0.053 | 0.380 | 0.108 | 0.072 |
| NEO (ne) | **0.146** | **0.015** | -0.029 | 0.630 | **-0.121** | **0.044** | 0.081 | 0.179 | -0.039 | 0.519 |
| NEO (ex) | -0.114 | 0.057 | **0.152** | **0.011** | -0.053 | 0.378 | 0.013 | 0.828 | -0.018 | 0.763 |
| NEO (op) | -0.060 | 0.315 | 0.094 | 0.116 | -0.081 | 0.177 | 0.097 | 0.107 | **-0.125** | **0.037** |
| NEO (ag) | -0.010 | 0.873 | -0.078 | 0.195 | -0.010 | 0.869 | 0.025 | 0.678 | 0.037 | 0.543 |
| NEO (co) | -0.111 | 0.063 | 0.024 | 0.687 | 0.049 | 0.413 | -0.079 | 0.190 | 0.066 | 0.274 |
| MDQ | 0.032 | 0.600 | 0.076 | 0.208 | **-0.123** | **0.040** | -0.076 | 0.207 | -0.005 | 0.938 |
| QIDS | 0.076 | 0.204 | -0.031 | 0.607 | 0.004 | 0.948 | 0.052 | 0.381 | -0.045 | 0.456 |
| STAI (pre) | 0.077 | 0.199 | -0.018 | 0.760 | 0.059 | 0.324 | -0.018 | 0.760 | 0.042 | 0.487 |
| CHRTP | 0.072 | 0.228 | -0.007 | 0.906 | -0.004 | 0.942 | 0.014 | 0.814 | -0.110 | 0.066 |
| SCQ | -0.087 | 0.157 | -0.022 | 0.724 | 0.012 | 0.840 | -0.034 | 0.585 | 0.025 | 0.686 |
| SHAPS (dics) | -0.027 | 0.652 | -0.057 | 0.338 | 0.012 | 0.844 | 0.024 | 0.687 | 0.022 | 0.716 |
| SHAPS (cons) | -0.029 | 0.624 | -0.058 | 0.334 | 0.020 | 0.733 | 0.006 | 0.917 | -0.011 | 0.850 |
| SAS (total) | 0.064 | 0.305 | 0.001 | 0.988 | -0.058 | 0.349 | -0.046 | 0.463 | 0.060 | 0.334 |
| SAS (mean) | 0.072 | 0.247 | -0.026 | 0.681 | -0.058 | 0.351 | 0.008 | 0.893 | 0.036 | 0.563 |
| SAPAS | 0.077 | 0.202 | 0.061 | 0.310 | -0.104 | 0.084 | 0.045 | 0.449 | -0.060 | 0.316 |
| AAQ | -0.006 | 0.918 | 0.046 | 0.441 | -0.067 | 0.265 | -0.090 | 0.134 | 0.037 | 0.544 |

| | | | | | | | | | | |
|---|---|---|---|---|---|---|---|---|---|---|
| CTQ (ea) | -0.006 | 0.917 | -0.058 | 0.331 | -0.001 | 0.988 | -0.073 | 0.222 | -0.014 | 0.814 |
| CTQ (en) | -0.074 | 0.215 | -0.055 | 0.363 | -0.023 | 0.702 | 0.005 | 0.938 | -0.026 | 0.661 |
| CTQ (pa) | -0.030 | 0.617 | -0.009 | 0.883 | 0.060 | 0.320 | -0.088 | 0.143 | 0.057 | 0.342 |
| CTQ (pn) | 0.019 | 0.757 | -0.002 | 0.971 | 0.069 | 0.248 | -0.069 | 0.251 | 0.016 | 0.794 |
| CTQ (sa) | 0.102 | 0.089 | 0.020 | 0.734 | 0.039 | 0.516 | **-0.124** | **0.039** | 0.029 | 0.625 |

**Table S2. The prediction performance between transformed features from all five common components and clinical measures for MDDs.** We used the projected common feature from all the five common FC components to train a multiple linear regression. The Pearson's correlation coefficients between predictive clinical scores and true scores of were shown. All *p* values were corrected by Bonferroni method. The significant one was bolded.

| | *r* | *p* |
|---|---|---|
| MASQ (gd) | 0.182 | 0.075 |
| **MASQ (ad)** | **0.209** | **0.014** |
| MASQ (aa) | 0.186 | 0.059 |
| **NEO (ne)** | **0.217** | **0.009** |
| **NEO (ex)** | **0.208** | **0.017** |
| **NEO (op)** | **0.202** | **0.024** |
| NEO (ag) | 0.090 | N.S. |
| NEO (co) | 0.163 | 0.205 |
| MDQ | 0.167 | 0.179 |
| QIDS | 0.110 | N.S. |
| STAI (pre) | 0.104 | N.S. |
| CHRTP | 0.138 | 0.724 |
| SCQ | 0.100 | N.S. |
| SHAPS (dics) | 0.070 | N.S. |
| SHAPS (cons) | 0.068 | N.S. |
| SAS (total) | 0.118 | N.S. |
| SAS (mean) | 0.109 | N.S. |
| SAPAS | 0.159 | 0.265 |
| AAQ | 0.128 | N.S. |
| CTQ (ea) | 0.099 | N.S. |
| CTQ (en) | 0.092 | N.S. |
| CTQ (pa) | 0.120 | N.S. |
| CTQ (pn) | 0.097 | N.S. |
| CTQ (sa) | 0.162 | 0.230 |

**Table S3. Summary of time-by-FC interaction effects in sertraline arm.** The important ROI-ROI FCs were identified for analysis based on the regression model specific to sertraline. All FCs with non-zero weight were selected. Connection label was categorized into high or low by above or below the median connectivity of all patients. Then, label of the group with better sertraline response across different weeks was shown. After FDR correction for statistical comparisons of all predictive FCs, only the significant ($p_{fdr} < 0.05$) time-by-FC interaction effects to $HAMD_{17}$ change with sertraline treatment were shown.

**Sertraline**

| **ROI1** | **ROI2** | ***p* value** | **Connection label** | **Location** |
|---|---|---|---|---|
| Supramarginal (R) | Postcentral (L) | 0.002 | High | 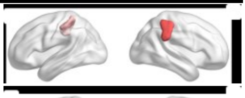 |
| Postcentral (R) | Middle Temporal (L) | 0.013 | High | 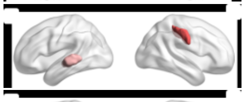 |
| Inferior Parietal (R) | Superior Temporal (R) | 0.013 | High | 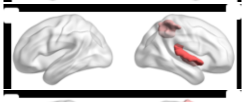 |
| Superior Frontal (R) | Rolandic Operculum (R) | 0.013 | High | 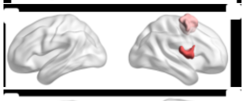 |
| Temporal Pole (R) | Insula (R) | 0.013 | Low | 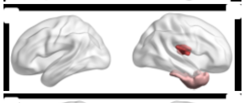 |
| Angular (R) | Middle Occipital (R) | 0.013 | High | 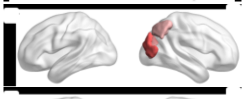 |
| Middle Frontal (R) | Inferior Temporal (R) | 0.013 | High | 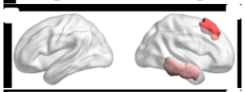 |
| Posterior Cingulate (R) | Precentral (L) | 0.013 | Low | 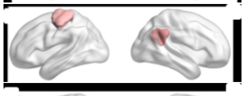 |
| Inferior Temporal (L) | Cuneus (L) | 0.016 | High | 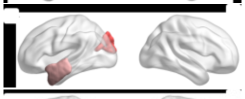 |
| Posterior Cingulate (R) | Inferior Temporal (L) | 0.033 | High | 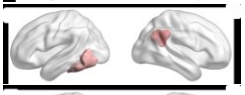 |
| Middle Cingulate (R) | Inferior Parietal (L) | 0.035 | High | 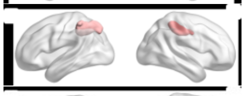 |
| Temporal Pole (R) | Inferior Temporal (L) | 0.035 | High | 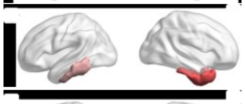 |
| Precuneus (R) | Angular (R) | 0.038 | High | 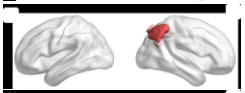 |
| Inferior Parietal (R) | Postcentral (L) | 0.043 | High | 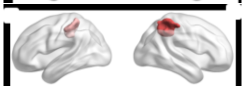 |

| | | | | |
|---|---|---|---|---|
| Posterior Cingulate (R) | Medial Orbitofrontal (L) | 0.043 | Low | 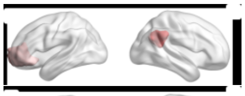 |
| Postcentral (L) | Lingual (L) | 0.043 | Low | 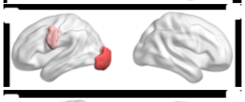 |
| Anterior Cingulate (R) | Supramarginal (L) | 0.049 | High | 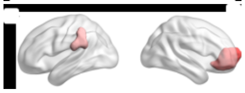 |

**Table S4. Summary of time-by-FC interaction effects in placebo arm.** The important ROI-ROI FCs were identified for analysis based on the regression model specific to placebo. All FCs with non-zero weight were selected. Connection label was categorized into high or low by above or below the median connectivity of all patients. Then, label of the group with better placebo response across different weeks was shown. After FDR correction for statistical comparisons of all predictive FCs, only the significant ($p_{fdr} < 0.05$) time-by-FC interaction effects to $HAMD_{17}$ change with placebo treatment were shown.

**Placebo**

| ROI1 | ROI2 | *p* value | Connection label | Location |
|---|---|---|---|---|
| Superior Temporal (L) | Middle Occipital (L) | 2.5E-07 | Low | 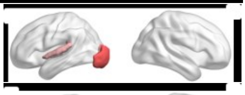 |
| Precentral (R) | Middle Occipital (L) | 3.2E-04 | Low | 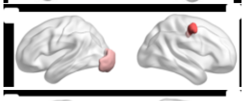 |
| Postcentral (L) | Middle Occipital (L) | 3.2E-04 | Low | 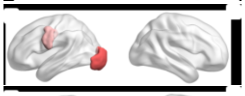 |
| Superior Temporal (R) | Middle Occipital (L) | 0.001 | Low | 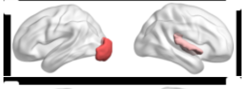 |
| Superior Temporal (L) | Middle Occipital (L) | 0.002 | Low | 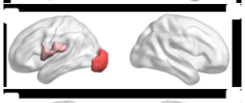 |
| Middle Frontal (R) | Middle Frontal (R) | 0.003 | High | 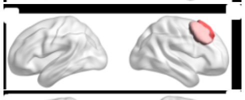 |
| Postcentral (R) | Middle Occipital (L) | 0.003 | Low | 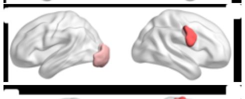 |
| Postcentral (R) | Middle Occipital (L) | 0.005 | Low | 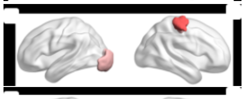 |
| Middle Frontal (L) | Inferior Temporal (L) | 0.005 | High | 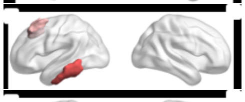 |
| Middle Frontal (L) | Middle Temporal (L) | 0.016 | Low | 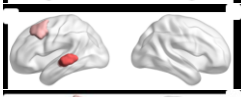 |
| Supramarginal (L) | Superior Parietal (L) | 0.028 | High | 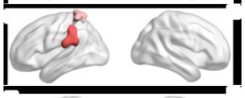 |
| Middle Frontal (R) | Posterior Cingulate (L) | 0.030 | Low | 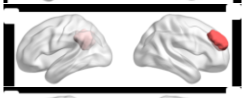 |
| Anterior Cingulate (R) | Posterior Cingulate (L) | 0.030 | Low | 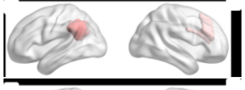 |
| Superior Temporal (R) | Lingual (L) | 0.030 | Low | 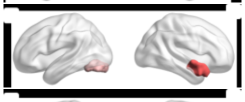 |
| Inferior Frontal (R) | Insula (R) | 0.030 | Low | 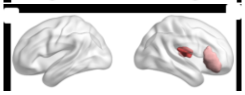 |

| | | | | |
|---|---|---|---|---|
| Inferior Frontal (L) | Inferior Temporal (L) | 0.030 | High | 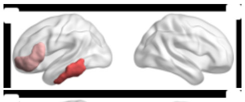 |
| Orbital Middle Frontal (R) | Supramarginal (R) | 0.032 | Low | 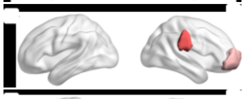 |
| Anterior Cingulate (R) | Supramarginal (R) | 0.032 | Low | 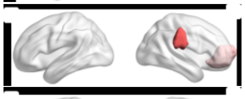 |
| Superior Temporal (R) | Lingual (L) | 0.049 | Low | 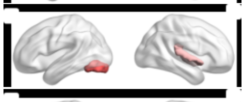 |
| Inferior Occipital (R) | Postcentral (L) | 0.049 | Low | 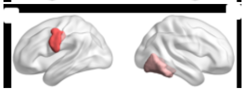 |
